# BRAVEHEART: Open-source software for automated electrocardiographic and vectorcardiographic analysis

**DOI:** 10.1101/2023.05.17.23290060

**Authors:** Hans Friedrich Stabenau, Jonathan W. Waks

## Abstract

**Background and Objectives:** Electrocardiographic (ECG) and vectorcardiographic (VCG) analyses are used to diagnose current cardiovascular disease and for risk stratification for future adverse cardiovascular events. With increasing use of digital ECGs, research into novel ECG/VCG parameters has increased, but widespread computer-based ECG/VCG analysis is limited because there are no currently available, open-source, and easily customizable software packages designed for automated and reproducible analysis.

**Methods and Results:** We present BRAVEHEART, an open-source, modular, customizable, and easy to use software package implemented in the MATLAB programming language, for scientific analysis of standard 12-lead ECGs acquired in a digital format. BRAVEHEART accepts a wide variety of digital ECG formats and provides complete and automatic ECG/VCG processing with signal filtering to remove high- and low-frequency artifact, non-dominant beat identification and removal, accurate fiducial point annotation, VCG construction, median beat construction, customizable measurements on median beats, and output of measurements and results in numeric and graphical formats.

**Conclusions:** The BRAVEHEART software package provides easily customizable scientific analysis of ECGs and VCGs. We hope that making BRAVEHART available will allow other researchers to further the field of EVG/VCG analysis without having to spend significant time and resources developing their own ECG/VCG analysis software and will improve the reproducibility of future studies. Source code, compiled executables, and a detailed user guide can be found at http://github.com/BIVectors/BRAVEHEART. The source code is distributed under the GNU General Public License version 3.

## 1. Introduction

The electrocardiogram (ECG) is a critical tool for clinical cardiovascular medicine and cardiovascular medical research. ECG analysis is used to diagnose existing cardiovascular disease and for risk stratification of future cardiovascular events, both in clinical practice and for research purposes. Although historically ECGs have been read by trained physicians on paper printouts, as medicine becomes more computerized, there has been increasing use of computerized ECG annotation and interpretation [1]. With increased availability of digitized ECGs there is also increasing interest in measurements which require additional signal processing of the standard 12-lead ECG and which cannot be obtained by visual inspection of the standard 12-ECG alone.

Computerization and digitization of ECGs have also allowed widespread analysis of vectorcardiograms (VCGs), where the electrical activity of the heart is visualized in 3 dimensions, and associated vectorcardiographic measurements [2]. These measures, which include the spatial ventricular gradient (SVG) [3], sum absolute QRST integral (SAI QRST) [4], QRST angle [5], total cosine R to T (TCRT) [6], and quantification of vectorcardiographic loop morphology [7], have been helpful in diagnosing current cardiovascular disease and predicting future adverse cardiovascular events across multiple studies with diverse patient populations.

Although clinically promising, widespread study of these and other VCG measurements has been limited because they require additional computer processing and specialized software. Previously described software includes the ECGlib software library and ECGlab graphical user interface (GUI) [8, 9], the Leiden ECG analysis and decomposition software (LEADS) software package [10], and other programs [11, 12]. However, these programs are not publicly available [10, 11, 9], and available open-source software packages [8, 12, 13, 14] are libraries for specific parts of ECG/VCG processing, such as R peak detection, dominant beat labeling, or fiducial point annotation, rather than full programs that allow complete processing of ECG data from file to measurements.

ECG/VCG research therefore usually requires investigators to create custom software which can be time consuming to develop and test, and which makes reproducible measurement difficult because variations in filtering, signal baseline definition, and signal processing preclude the direct comparison of measurements. Reproducible analysis is also a special priority as the reproducibility of results is of growing concern in scientific research [15]. We therefore set out to develop open-source, easily customizable software designed to support ECG/VCG research with high-throughput, automated, and reproducible ECG and VCG analysis with the Beth Israel Analysis of Vectors of the Heart (BRAVEHEART) software library.

BRAVEHEART addresses the limitations of existing software as a modular software library meant to perform reproducible annotations and ECG and VCG measurements for scientific analysis on standard 12-lead ECGs acquired in a digital format. The design goals of BRAVEHEART include:

- A customizable, modular, and easily extensible design makes it simple to add new measurements or ECG formats as needed.
- Signal filtering and baseline correction performed in a physiologically appropriate way.
- Automated operation that is robust to non-dominant beats (premature ventricular contractions [PVCs], aberration, ventricular pacing, etc.), artifact, and noise.
- Reproducible ECG/VCG annotation, signal segmentation, and measurements.
- Automatic and efficient batch processing of ECGs with parallel processing capability.
- Open source MATLAB (MathWorks, Natick, MA) code as well as compiled executable code for researchers without access to MATLAB.

## 2. Methods/Software Description

BRAVEHEART is a software package which performs automated ECG/VCG analysis for research purposes. It takes a digital 12-lead ECG signal in a variety of formats, and provides automatic signal filtering, QRST fiducial point annotation, median beat alignment and construction, and measurement of a variety of ECG/VCG parameters on the median beat. It is also designed to be easily customizable. BRAVEHEART currently runs on MATLAB versions above R2022a. Matlab source code, compiled executables for Windows and Mac operating systems, and a detailed user guide can be found at http://github.com/BIVectors/BRAVEHEART. The source code is distributed under the General Public License (GPL) version 3.

### 2.1. BRAVEHEART Overview

BRAVEHEART ECG processing proceeds via the following steps:

1. Reading in the digital ECG in a variety of formats.
2. Wavelet-based filtering for high-frequency and low-frequency (baseline wander) noise removal.
3. VCG construction.
4. Peak thresholding for heart rate (HR) estimation and locating QRS complexes.
5. Baseline offset correction of the VCG leads.
6. Median filtering for estimation of QRS width and automatic pacemaker spike detection and removal.
7. Preliminary heuristic fiducial point (QRS onset [*Q*_on_], QRS offset [*Q*_off_], and T wave offset [*T*_off_]) annotation.
8. Non-dominant QRST morphology (PVC, ventricular pacing, or aberration) detection: non-dominant beats can be analyzed separately or automatically excluded from subsequent analyses.
9. Outlier beat detection: outlier beats can be automatically excluded.
10. Median beat VCG construction.
11. Median beat fiducial point (*Q*_on_, *Q*_off_, and *T*_off_) annotation using a custom neural network (NN).
12. VCG signal and annotation quality assessment.
13. Graphical display and analysis of the VCG median beat.
14. Export of ECG/VCG measurement data to an external file for analysis.

### 2.2. Reading Digital ECG Formats

BRAVEHEART is able to read a variety of common digital ECG formats, including extensible markup language (XML), from a variety of ECG recording system manufacturers such as General Electric (GE), Philips, and Mortara Instruments. HL7 XML is also supported. Other currently readable formats include Digital Imaging and Communications in Medicine (DICOM), International Society for Holter and Noninvasive Electrocardiology (ISHNE) .ecg files, GE Marquee .mrq files, GE Prucka format .txt files, and unformatted numeric lead data in text files. Each file format has its own load module, and it is therefore easy to integrate new file formats as needed.

### 2.3. ECG Signal Filtering

Once ECG signals are loaded, wavelet-based filtering is performed to remove noise/artifacts which can hamper ECG segmentation and processing. For high-pass filtering of each ECG lead, a discrete wavelet decomposition is computed. The approximated baseline signal is then reconstructed using wavelet decomposition levels below a configurable bandpass which is nominally set to 0.24 Hz. This low frequency signal which approximates baseline wander and respiration effects without influencing the main frequencies contained in the QRST complex (0.5-40 Hz) [16], is then subtracted from the original signal.

Low-pass filtering is achieved for each lead using a maximal overlap discrete wavelet decomposition using a level which is nominally set to the 62.5 Hz bandpass with a soft thresholding scheme to remove high frequency noise above this threshold. For both high- and low-pass filtering, the wavelet and level used as a frequency cutoff are adjustable by the user, although the nominal settings of high-pass at 0.24 Hz and low-pass of 62.5 Hz work well for most applications. Certain ECGs with significant baseline wander or high frequency artifact may require adjustment of these settings. Filtering can also be disabled if needed for specific applications. An example of ECG filtering and baseline wander removal using this methodology is shown in Figure 1.

**Figure 1:**
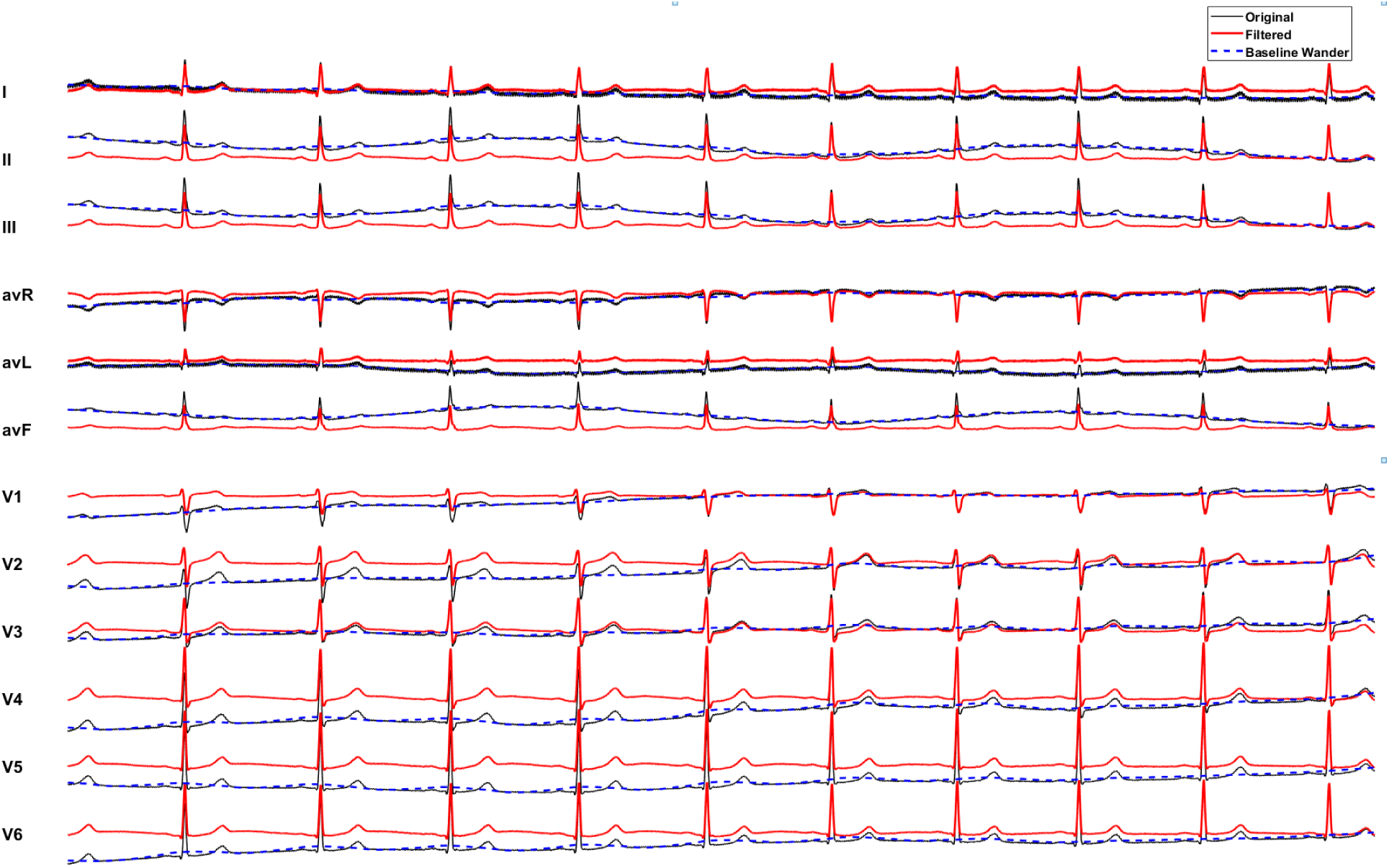
Example of utilizing wavelet decomposition to remove significant low-frequency artifact from an ECG with a sampling rate of 500 Hz. The original ECG signal is in black and the filtered signal is in red. In this case the significant baseline wander, as illustrated by the dashed blue line, is estimated and then subtracted out of the final signal so that the baseline is flat. In this example, modest high-frequency noise is also reduced.

### 2.4. Vectorcardiogram (VCG) Construction

A 3 ***×*** *n* VCG matrix (**V**) with orthogonal X, Y, and Z leads as its rows is constructed from a *n **×*** 8 ECG matrix (**E**) with the 8 independent ECG leads (I, II, V1-V6) as its columns using a 3 ***×*** 8 transformation matrix (**M**):

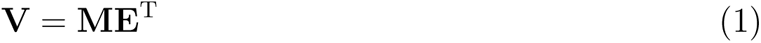

The Kors transformation matrix [17] is nominally used for **M**, although other transformation matrices [2] (such as the inverse Dower transformation [18]) can be substituted. The vector magnitude (VM) lead is then constructed by taking the Euclidean norm of the VCG X, Y, and Z leads:

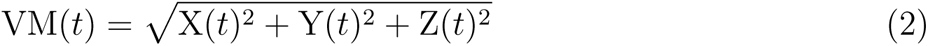

The VM signal is used for all subsequent annotations as it represents the global electrical activation of the heart as a combination of all 8 independent leads [19], and it is always positive which simplifies signal processing and annotation.

### 2.5. QRS Peak Thresholding

QRS complexes are detected in the VM lead by looking for peaks nominally in the top 5% of values, subject to a maximum HR constraint which helps in cases where there is significant fractionation of the QRS complex or very large amplitude T waves. The threshold parameter and maximum HR constraint may be manually adjusted in order to process ECGs that feature high-amplitude measurement artifacts, very peaked T waves, large differences in amplitude between native QRS complexes and PVCs, or paced QRS complexes. Peak thresholding is illustrated in Figure 2.

**Figure 2:**
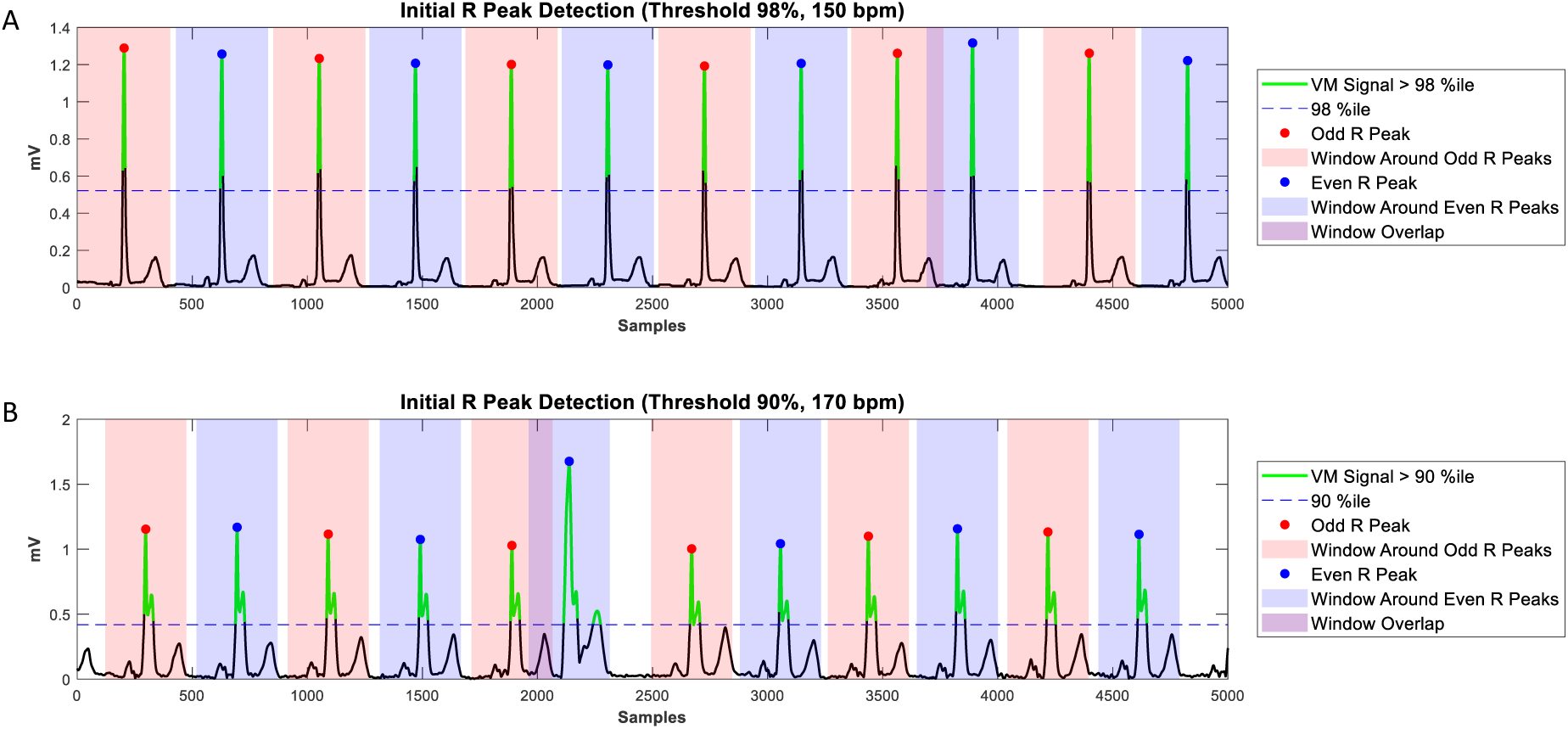
Example of VM lead peak thresholding with HR constraints. **A:** Parts of the signal which are above the 98th percentile are highlighted in green. R peaks are chosen as the dominant peak within a window (odd: red, even: blue, with width set by a chosen maximal HR). In this example the R peaks are significantly larger than the T waves and there are not any significant extra peaks in the QRS complex, and R peak detection is therefore straightforward. **B:** In this example there are 2 peaks in each QRS complex and a PVC. The threshold was artificially lowered below what would nominally be chosen for illustrative purposes. Even with a lower than normal threshold of 90%, only a single peak is found for each QRST complex, and the second minor QRS peak and the PVC T wave are ignored. Had the PVC been more tightly coupled the maximum HR constraint could be increased. **Abbreviations**: PVC - premature ventricular contraction, VM - vector magnitude, HR - heart rate.

### 2.6. Baseline Offset Correction of the Vectorcardiogram

ECG machines usually assign the zero voltage reference to a point that is not necessarily the physiological zero voltage. This may be computationally advantageous during ECG signal acquisition using an ECG machine where visual inspection of the baseline rather than exact voltage measurement relative to zero voltage is performed in most cases and for most clinical purposes. However, when measurements are being made on ECG leads, especially measurements involving area under the QRST complex, an inaccurate baseline zero voltage reference can lead to large errors in these measurements (See Supplemental Figure 1). Filtering can introduce baseline offsets as well. Therefore, in order to ensure accurate measurements, a method of finding the physiologic zero voltage reference after filtering is needed.

During the TP interval, after ventricular repolarization is complete and before start of atrial depolarization (in sinus rhythm), the heart is electrically silent, and therefore the voltage in all ECG leads should approximate 0 mV. While each ECG lead does undergo baseline wander correction during high-pass filtering as noted above, and this can remove large constant (0 Hz) shifts, there is often still a residual baseline offset which can be due to noise, respiration, or amplifier drift. The high-pass filter does not use any physiologic information and has no way of ensuring that the TP segment of the signal equals 0 mV. When these offset ECG leads are then projected onto X, Y, and Z coordinates, the residual offsets can add up to an even larger baseline offset [20, 21].

In order to optimize the VCG for quantitative measurements such as area and angle measurements, it is essential to zero the ECG lead baselines in a physiologically meaningful way by adding a constant offset so that the TP segment approximates 0 mV voltage. This re-zeroing process can be performed at any time before or after transformation of the ECG into a VCG, but doing so after VCG construction is advantageous as fewer leads need to be corrected (3 vs 8), and the ECG baseline offsets have no effect on the transformation from ECG to VCG other than introducing a new offset in the transformed signals (matrix multiplication is distributive for constants).

For each lead, this offset is determined by the following procedure: First, the signal is smoothed with a 4th order Savitzky-Golay filter with window size equal to 10% of the average cardiac cycle length which has the effect of smoothing out sharp features of the ECG signal. Next, the set of points corresponding to flat regions of the ECG signal is determined by finding the set of points where the slope of the filtered signal is less than 2% of the maximum slope. Finally, for each lead, the median of this set of points is determined, which is then the offset by which the lead is subsequently shifted. This baseline offset procedure is robust to noisy signals and atrial fibrillation, and functions as long as the HR is not so tachycardic that there is no appreciable TP segment (or TQ segment in atrial fibrillation) which would render the ECG difficult to quantitatively analyze for other reasons. An example of baseline correction is shown in Figure 3.

**Figure 3:**
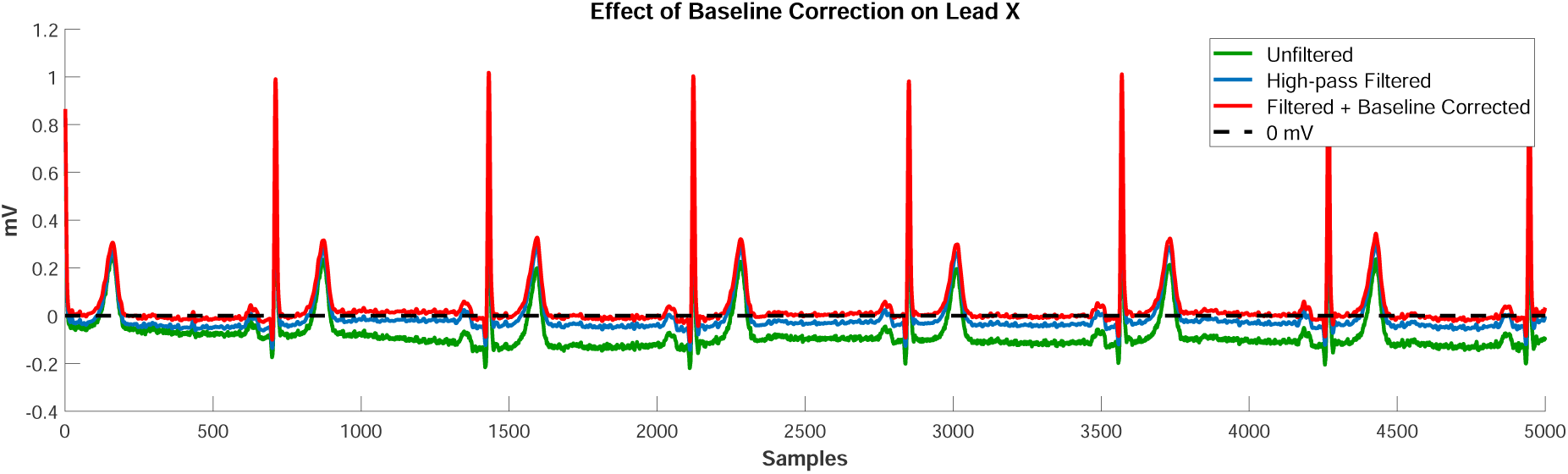
Example of baseline correction after filtering. The dashed black line corresponds to 0 mV. The green signal is the raw unfiltered ECG signal (lead X) with 0 mV voltage assigned by the ECG recording system. The blue signal is lead X after high-pass filtering; most of the baseline offset is removed by filtering, but the TP segment is not appropriately set to zero voltage. The red signal is lead X after additional baseline correction as described in the methods. After baseline correction the TP segment lead X now approximates 0 mV.

### 2.7. Median Filtering for Pacemaker Spike Detection

For each peak detected in the VM lead, pacemaker spikes are identified (if present) with the following procedure: first, the signal is smoothed with a 4 ms median filter. The start and end of each R peak previously detected in the VM signal (see Section 2.5) is annotated by marching out from the peak forwards and backwards and marking where the signal first crosses an adjustable threshold which is nominally set at 20% of the peak height.

Candidate QRS peaks that are less than a certain threshold in width (default: 20 ms) are marked as pacing spikes, as these spikes occur over time intervals that are not physiologic for a QRS complex. If pacing spikes are found they are simply ignored and the QRS-finding procedure is repeated. The HR is then estimated as the mean distance between R peaks (RR intervals) over the entire ECG.

### 2.8. Heuristic First Pass VCG Fiducial Point Annotation

It has been shown that performing calculations on median beats is preferable to calculating a quantity on each beat individually and then finding the mean or median of measurements [19, 22]. Accurate annotation of each individual QRST complex is therefore important primarily to allow accurate alignment of each individual beat to create an accurate median beat which will be used for all subsequent measurements. For this reason, it is less critical that each individual beat be perfectly annotated, as small errors in fidicual point annotation do not preclude accurate median beat creation which relies primarily on aligning the major features of the QRS complex/peaks rather than aligning beats based on the exact location of fiducial points.

To allow rapid computation of fiducial points with a simple way of visualizing the annotation algorithm and troubleshooting problematic ECGs or ECGs with atypical QRST complexes, first pass QRST annotation utilizes standard signal processing methods on the VM lead. For each R peak that is present, the QRS width is first estimated (similarly to the previous step in Section 2.7) using a 40 ms median filter with an adjustable threshold nominally set at 20%. A QRS search window is then defined around each R peak. The adjustable QRS search window is set to be equal in duration to twice the estimated QRS width, centered around the R peak. The adjustable T wave search window starts by default 100 ms after the end of the located S-wave/*Q*_off_ and nominally extends forward to 45% of the mean RR interval. Rarely, it is necessary to adjust this percentage based on HR, QT interval, and QRST morphology. The onset and offset of the QRS complex are determined by an algorithm that is described in detail in the “Heuristic Annotation Details” section of the Supplemental Methods (Supplemental Section 1.1). Details on the search windows and an example of first pass annotation is shown in Figure 4.

**Figure 4:**
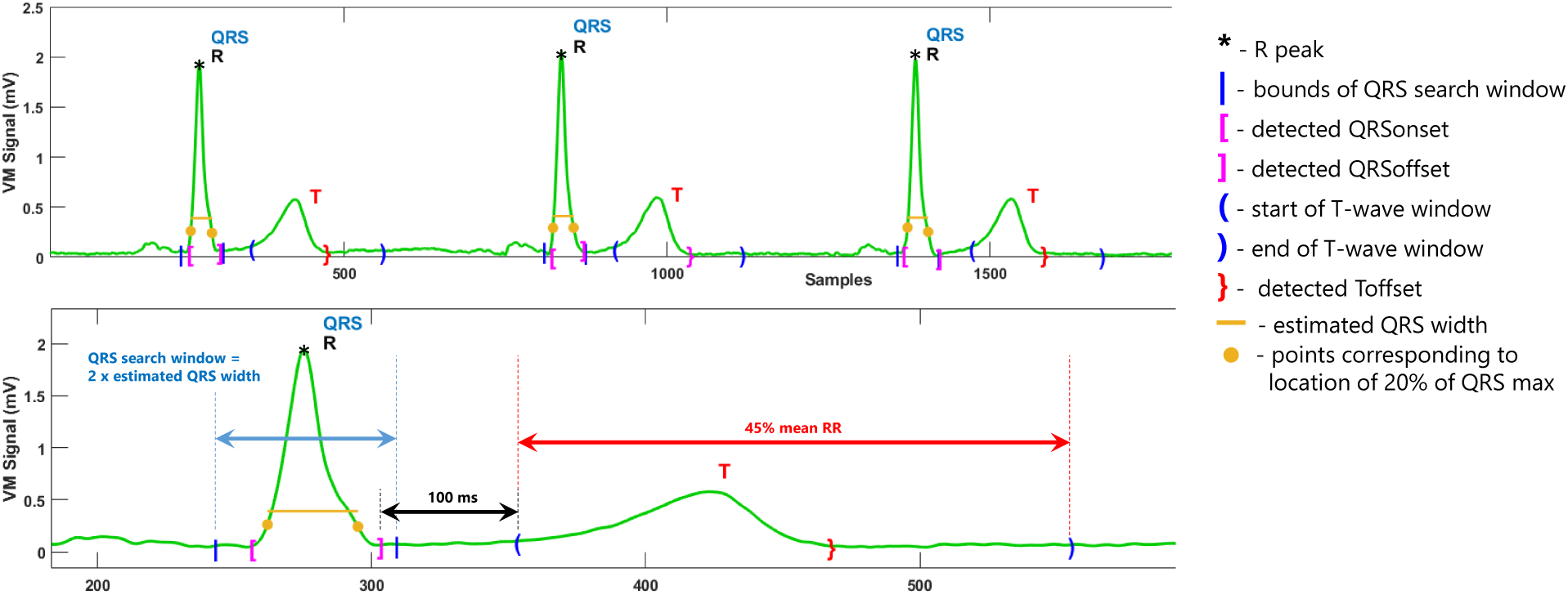
Example of first pass heuristic annotation. The upper panel shows 3 sequential beats, and the lower panel is zoomed in on beat 1. Search windows and the methodology for fiducial point detection are described in detail in the text. Using nominal settings, the QRS search window (blue arrow and blue *|* symbols) is set as 2 times the estimated QRS width (yellow bar) centered around the QRS peak. The T wave search window (red arrows and blue parentheses) nominally starts 100 ms after QRS offset, and extends forward 45% of the mean RR interval. These nominal parameters are adjustable.

T-wave offset detection can use baseline crossing, the tangent method [23], or a validated method that is more robust to abnormal T wave morphology that estimates minima of the T wave “energy” and which is more accurate than these other methods, especially in ECGs with low amplitude or notched T waves [24]. After first pass annotation, beats too close to the start or end of the ECG so that an entire QRST complex is not present are removed from analysis.

### 2.9. Non-dominant Beat (PVC, Pacing, and Aberration) Detection

PVCs or other non-dominant beat classes, which include intermittent QRS aberration or intermittent ventricular pacing, are detrimental to median beat construction, especially when frequent non-dominant beats are present. For example, in cases of ventricular bigeminy where there are equal numbers of PVCs and normal QRST complexes, unless PVCs are removed, the median QRST complex will be be a superposition of the PVC and normal QRST morphologies, with a morphology that is similar to neither (see Figure 5). For this reason it is also not always possible to simply compare individual beats to the median beat for non-dominant beat detection. BRAVEHEART automatically identifies and can remove non-dominant QRST morphologies. This process will identify any non-dominant QRST morphology including PVCs, intermittent ventricular pacing, and intermittent QRS aberration. All of these types of non-dominant QRST complexes are considered “PVCs” for purposes of the following method description.

**Figure 5:**
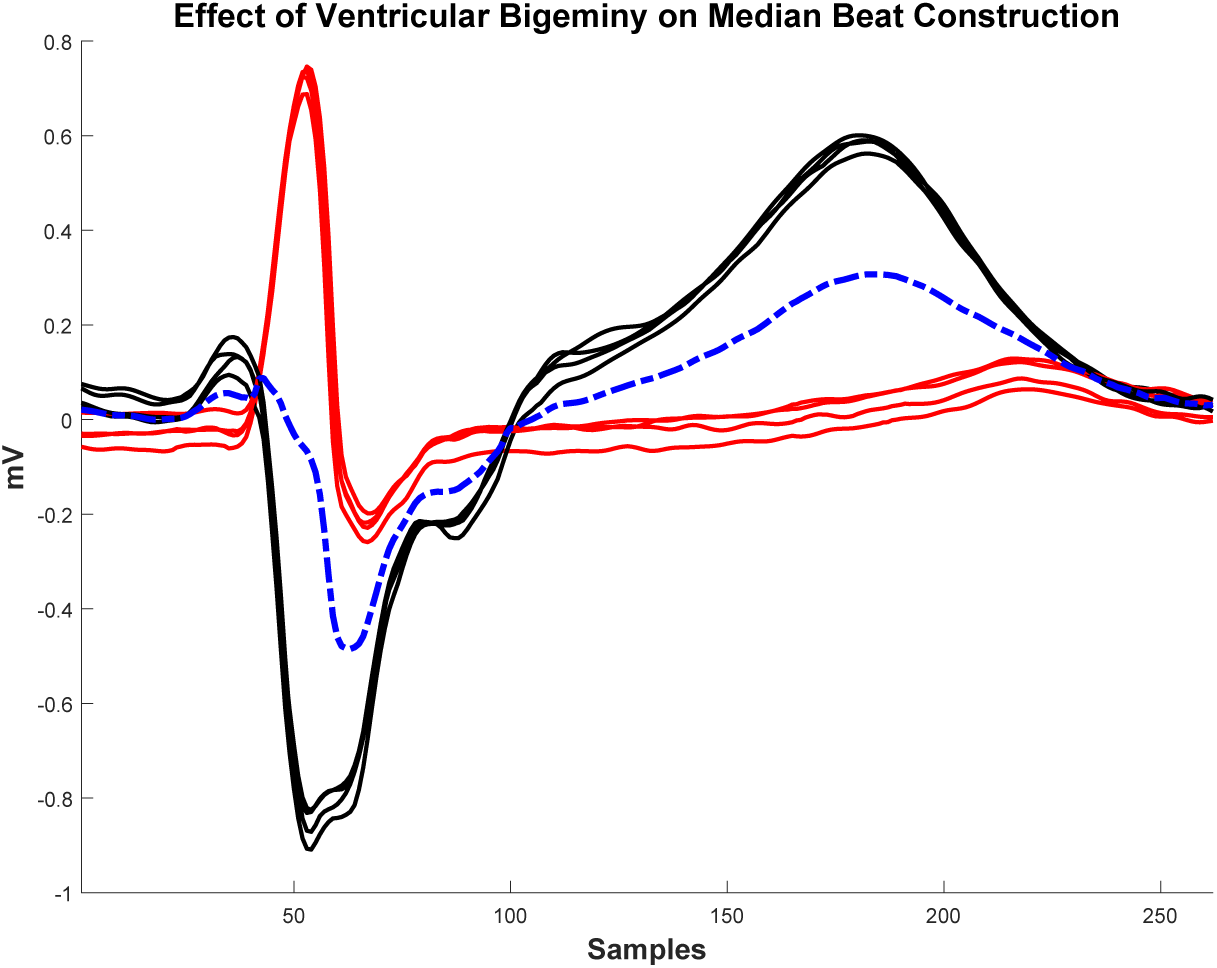
Effect of ventricular bigeminy on median beat construction. If ventricular bigeminy is present and the full ECG strip contains equal numbers of normal beats (red) and PVCs (black), the created median beat (dashed blue line) will be a superposition between the normal QRST complexes and PVC QRST complexes, and will result in incorrect measurements. **Abbreviations**: PVC - premature ventricular contraction.

BRAVEHEART PVC detection utilizes normalized cross correlation (NCC) and root mean squared error (RMSE) to perform a form of “template matching” between individual QRST complexes. A detailed description of the PVC detection algorithm is available in the “PVC Detection Algorithm Details” section of the Supplemental Methods (Supplemental Section 1.2), Figure 6, and Supplemental Figures 2 and 3.

**Figure 6:**
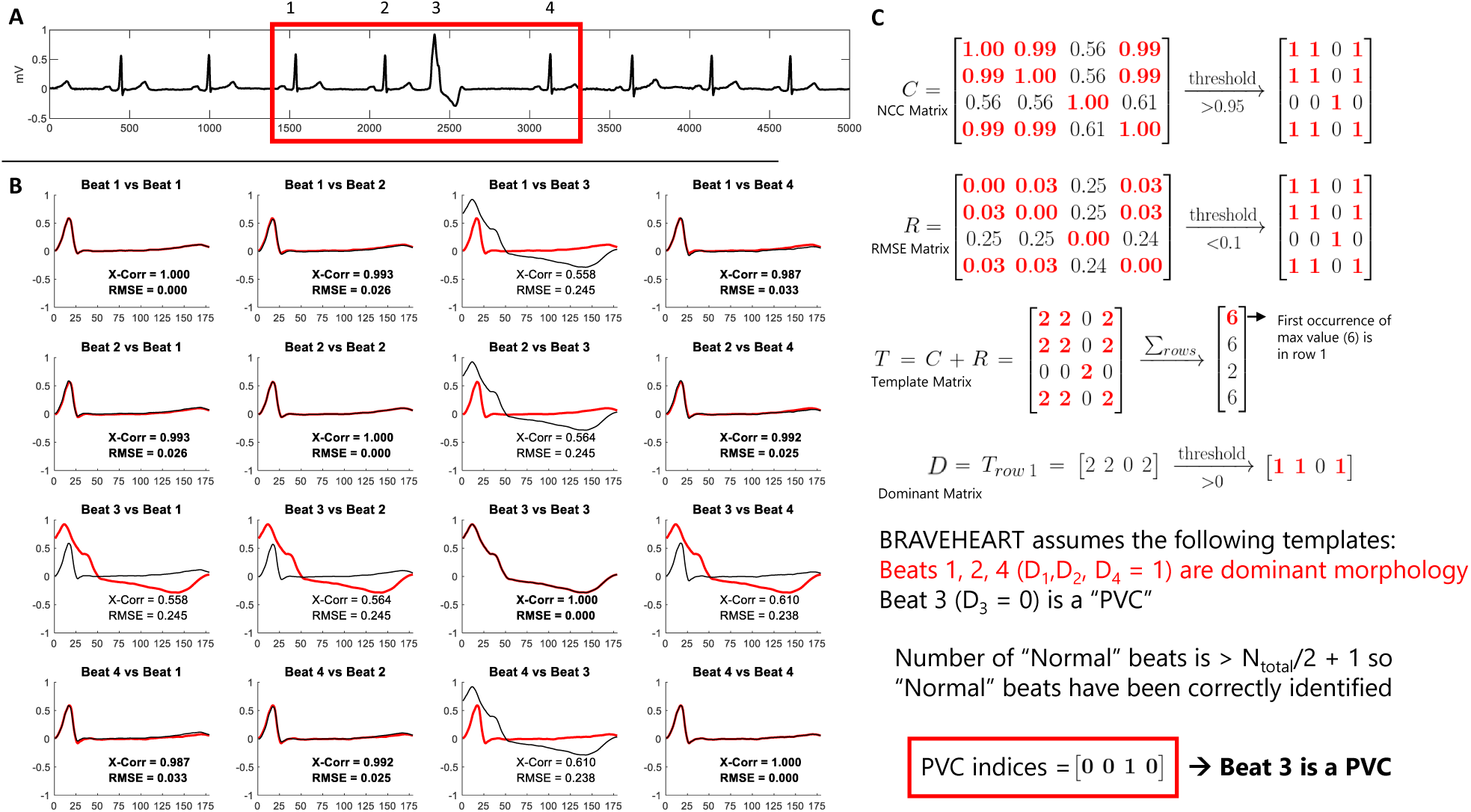
Example of how the BRAVEHEART PVC detection algorithm works on an ECG with a single PVC. **A:** A subset of 4 beats (3 sinus and 1 PVC) are analyzed for illustrative purposes, but the algorithm would have the same result if all 9 beats were analyzed. **B:** After trimming, each beat is aligned with the remaining beats, and normalized cross correlation (NCC) and root mean squared error (RMSE) are calculated. **C:** Values of NCC and RMSE are placed in matrices *C* and *R*, respectively, and then thresholds are applied as described in the text. The template matrix *T* is formed by adding *C* + *R* and summing the rows. The dominant matrix *D* is formed by taking the first row of *T* that has the maximum row sum value, and then performing additional thresholding. BRAVEHEART starts by assuming that values of 1 in *D* are the dominant morphology and values of 0 in *D* are PVCs. In this case, since the number of beats with dominant morphology is *> n/*2 + 1, the algorithm is complete and the indices of PVCs are found by taking values of 0 in matrix *D* (beat 3). **Abbreviations**: PVC - premature ventricular contraction.

Using a single lead for PVC detection may limit the sensitivity and specificity of PVC detection because PVCs can be similar in morphology to dominant, non-PVC QRST complexes in one ECG lead, while having dramatically different morphology in other leads. BRAVEHEART therefore performs the PVC identification algorithm on the 3 orthogonal X, Y, and Z leads separately. A beat is ultimately identified as a PVC if it is identified as a PVC in at least 2 of the 3 orthogonal leads. This also helps avoid artifact in a single lead triggering incorrect PVC identification. The beats identified as PVCs can be removed from subsequent analyses, including median beat construction. The software also has the ability to also keep PVCs and remove non-PVC QRST complexes should analysis of PVCs be desired.

#### 2.9.1. Non-dominant Beat Detection Performance

The performance of the BRAVEHEART PVC detector was assessed in a sample of 286 sequential ECGs obtained from persons with a history of PVCs, totaling 3,488 total beats and 508 PVCs/ventricularly paced/aberrant beats (14.6%). Not all ECGs contained PVCs. Ground truth was assigned by manual review of the ECGs and labeling beats as “dominant” or “PVC”. We excluded 26 beats where focal artifact/noise significantly distorted what was likely a normal beat to the degree that it was morphologically distinct enough that it would be reasonable to consider it a “PVC” and to remove it from the analysis.

BRAVHEART’s PVC detection algorithm was then used to process this ECG dataset with various values of NCC and RMSE. Summary statistics, including sensitivity, specificity, positive predictive value, negative predictive value, F1 score, and overall accuracy were recorded. Performance was further assessed using bootstrapping using 1,000 replications to obtain confidence intervals for these measurements. PVC detection performance is presented in the Results Section (Section 3.1).

### 2.10. Outlier Detection/Removal

After PVCs, paced beats, and aberrant beats are removed, remaining beats with artifact that preclude accurate fiducial point annotation are likewise detrimental to accurate median beat construction. Although most beats with enough artifact to significantly change their QRST morphology compared to the dominant beat morphology will be removed via PVC detection, some beats will retain the dominant QRST morphology but have enough artifact so that they cannot be aligned with other beats accurately. They may also introduce undesired noise into median beat construction. Noisy beats of the dominant morphology can also escape PVC detection if there is focal artifact towards the end of the T wave that makes annotation difficult or impossible because the PVC detection adjusts the ends of the QRST complexes to make them all equal length (see Supplemental Section 1.2).

To deal with this situation, BRAVEHEART has an algorithm to identify and remove outlier beats within the dominant QRST morphology. For each beat, the QR and RT intervals are computed. A modified Z-score (*Z_i_*) for each of these quantities is then computed [25]. Details of modified Z-score calculation can be found in the “Outlier Detection Using Modified Z–Scores” section of the Supplemental Methods (Supplemental Section 1.3).

Nominally, any beat where the absolute value of *Z_i_* for either the QR or RT interval is greater than 3.5 is marked as an outlier [25]. These outliers are nominally removed automatically. The modified Z-score threshold that reliably detects outliers depends on the total number of beats being compared and can be adjusted as needed; in cases of bradycardia, it may be necessary to slightly increase the threshold to avoid tagging acceptable beats as outliers [25]. It is also possible to automatically adjust the modified Z-score threshold based on HR (and the number of beats in the ECG) for this purpose.

### 2.11. Median VCG Construction

The *n* remaining beats, which should now be all of a single morphology and free of excessive noise/artifact are aligned to create a median beat which is subsequently used for all calculations and measurements. Although BRAVEHEART has the ability to align the individual QRST complexes based on their R peak locations, this method is, in general, less reliable as it is highly influenced by artifact and noise. To overcome this limitation and obtain more robust beat alignment, we calculate the “center of voltage” (CoV) of each of the remaining *n* QRS complexes in each lead in a manner analogous to calculating center of mass. For a given QRS complex, let *S_i_* be sample *i* within it. The CoV of this QRS complex is then given by:

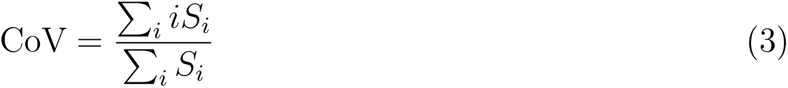

For each of the *n* QRST complexes, the CoVs are then used as the fiducial point for alignment. The signal duration incorporated into the median beat prior to the CoV is the maximum Q-CoV interval of all beats + 40 ms and the signal duration incorporated into the median beat after the CoV is the maximum CoV-T interval of all beats + 60 ms. The window is expanded like this so that small errors in the prior heuristic annotation will not result in cutting off the true *Q*_on_ or *T*_off_ before being passed into the final median beat annotation algorithm (see Section 2.12 below). Once all beats are the same duration and aligned properly, the median beat is obtained by computing the median voltage value on these aligned samples (see Figure 7). Median X, Y, and Z leads are computed. The median VM lead is then calculated from the median X, Y, and Z leads as previously described in Equation 2.

**Figure 7:**
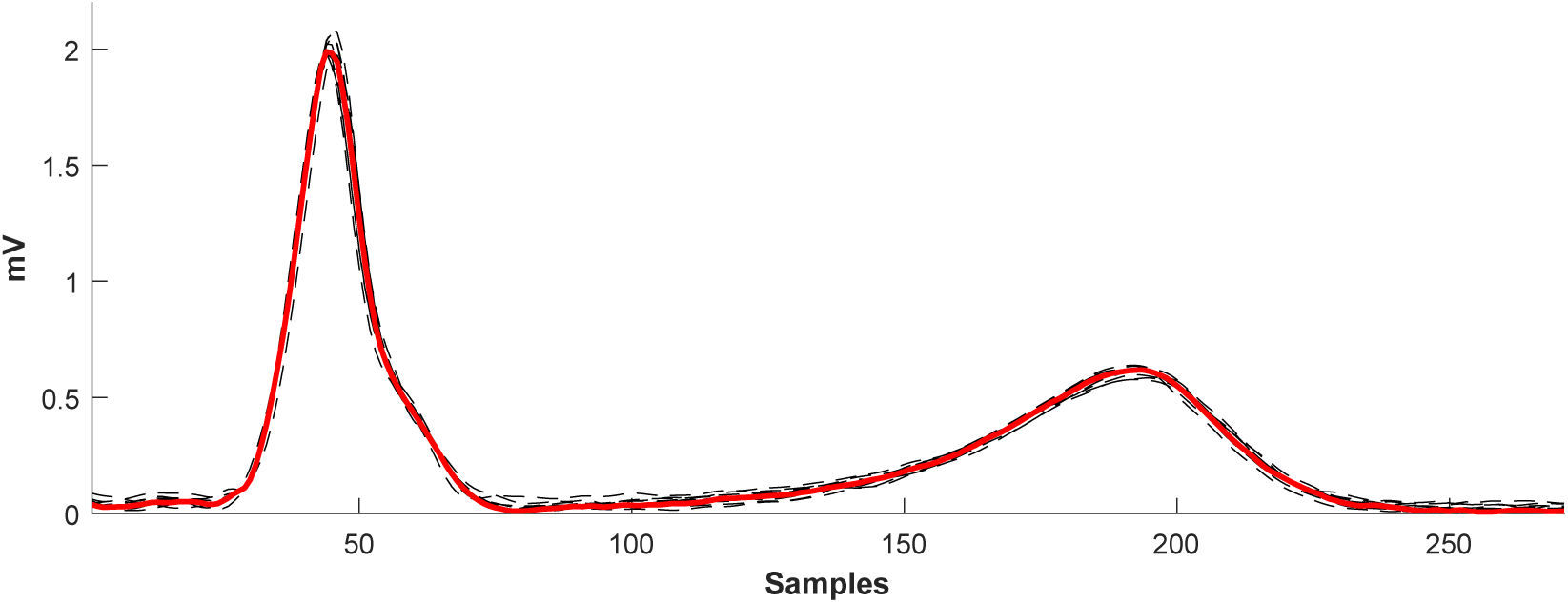
Example of median beat construction. Individual QRST complexes (black dashed lines) are aligned on their center of voltage, and the median of the aligned samples is calculated to obtain the median beat (red).

### 2.12. Median Beat Annotation

Accurate annotation of the median beat is critical for accurate measurement of ECG/VCG parameters. We found that the same heuristic methods for ECG annotation used in first pass annotation would often fail at annotating the median beat with enough accuracy for this purpose, especially in cases with atypical QRS complex morphology or when T waves were low amplitude or of abnormal morphology. We therefore created a custom bidirectional long short term memory neural network (NN) specifically for BRAVEHEART to accurately annotate fiducial points on VM median beat QRST complexes with a high degree of accuracy.

Details of NN architecture, training, and testing have previously been reported [26], and are also available in the Supplemental Methods Sections 1.4, 1.5, and 1.6. In brief, the NN takes in a variable length VM median beat signal and outputs probabilities of each sample of the signal being a “QRS complex”, “T wave”, or “other”. Transitions between “other” to “QRS complex” are taken as the location of *Q*_on_, transitions between “QRS complex” to “T wave” are taken as the location of *Q*_off_, and transitions between “T wave” to “other” are taken as the location of *T*_off_. See Figure 8 for an illustration of this process. In rare cases where the NN output results in multiple locations where the definition for one of the fiducial points was satisfied, logic based on the physiologic properties and order of QRST signals attempts to identify the correct fiducial point location.

**Figure 8:**
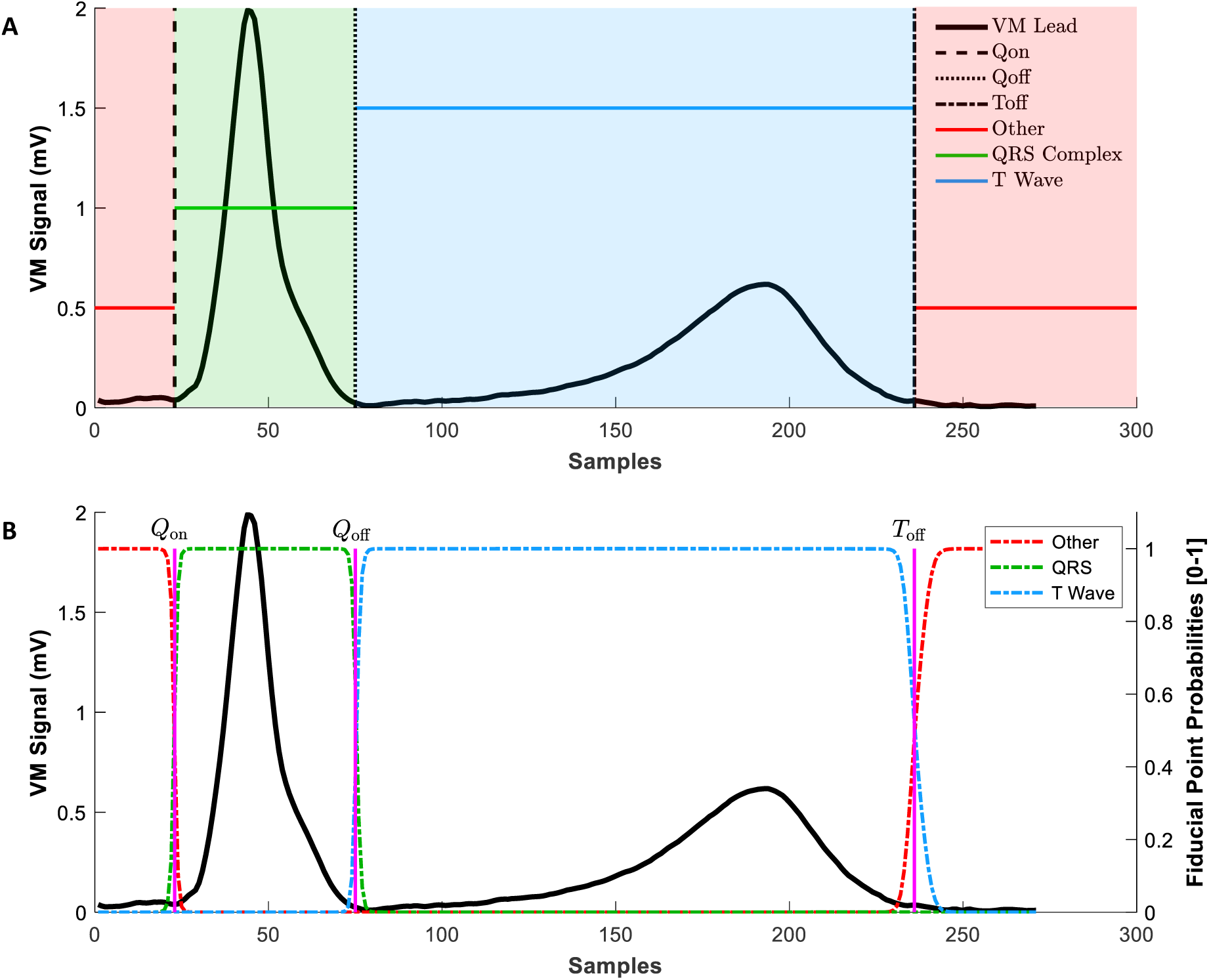
Signal processing and labeling for NN training/output. **A**: For NN training, 12-Lead ECGs were transformed to VCGs and the median VM beat was constructed (black signal). *Q*_on_, *Q*_off_, and *T*_off_ were annotated by a cardiac electrophysiologist as shown (black dashed, dotted, and dash-dotted lines, respectively). A categorical signal (red, green, and blue lines/shading) was constructed based on the location of *Q*_on_, *Q*_off_, and *T*_off_. The QRS complex was labeled between *Q*_on_ and *Q*_off_ (green). The T wave was labeled between *Q*_off_ and *T*_off_ (blue). The remainder of the signal was labeled as “other” (red). The labeled categorical signal was used for NN training/testing. See the text for details. **B**: Example of NN output. The probabilities of “QRS complex” (green), “T wave” (blue), and “other” (red) labels are shown relative to the median VM beat signal and the predicted fiducial points which are assigned at transitions between fiducial point probabilities as described in the text. **Abbreviations**: ECG - electrocardiogram, VCG - vectorcardiogram, VM - vector magnitude, NN - neural network, *Q*_on_ - QRS complex onset, *Q*_off_ - QRS complex end, *T*_off_ - T wave end.

#### 2.12.1. Median Beat Neural Network Annotation Performance

NN performance was assessed primarily based on calculating the mean and standard deviation of difference between NN predicted locations of *Q*_on_, *Q*_off_, and *T*_off_ and measured QRS duration and QT interval, and ground truth annotations performed by a board certified cardiac electrophysiologist (JWW). Given that there were 3 output classes, micro-averaged F1 was reported. As this was a multi-class labeling problem, the recall (specificity), precision (positive predictive value), and F1 (the harmonic mean of precision and recall) – see Supplemental Statistical Methods (Supplement Section 1.8) are all equal, and therefore only F1 was reported.

The included NN was tested on 189 median beats which were randomly allocated for testing during NN training. Given that BRAVEHEART users care most about the performance of the included weights and biases rather than overall theoretical model performance which has previously been described [26], we also validated NN performance on a second independent dataset of 200 sequentially obtained and manually annotated median beats. Details of NN performance is described in the Results Section 3.2.

### 2.13. ECG/VCG Parameter Calculations

BRAVEHEART has easily customizable modules that calculate a variety of ECG and VCG measurements on the median beat. Parameters include standard measurements such as HR, QRS duration, and QT interval, morphological measurements for each of the 16 leads such as R wave, S wave, and T wave magnitude (see Figure 9), and a variety of vectorcardiographic parameters including spatial QRS-T angle [5], TCRT [6], and peak and area vectors for the QRS complex, T wave, and entire QRST complex (spatial ventricular gradient [3]). VCG speed, loop coplanarity (how well the loops fit into a best fit plane), roundness (how circular vs oval), length, and area are also calculated. Further details and equations for calculated parameters are available in the “Equations” section of the Supplemental Methods and Supplemental Tables 1, 2, and and 3.

**Figure 9:**
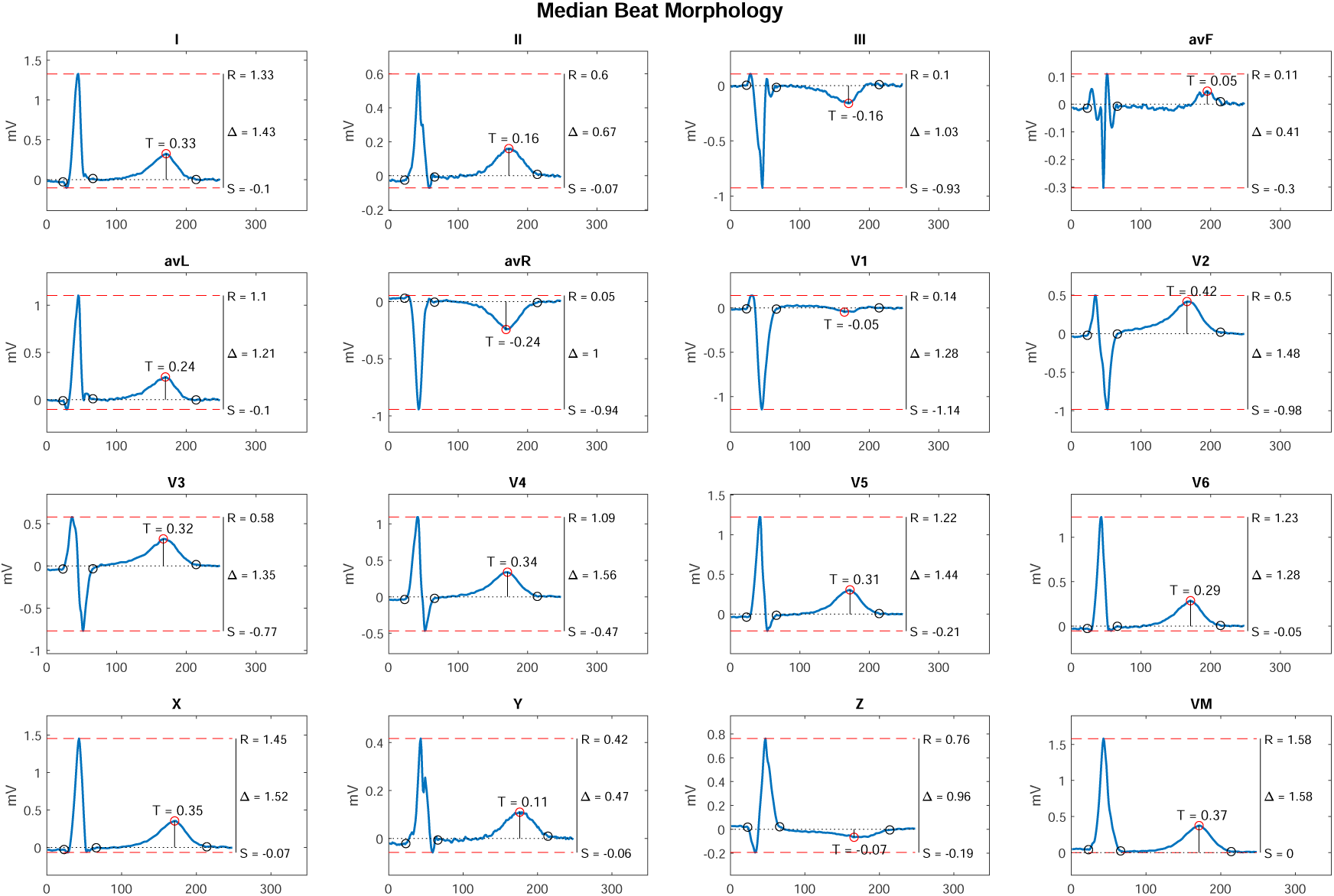
Lead morphology measurements including the magnitude of R, S, and T waves for all 16 leads are shown.

The software is designed to facilitate easy addition of new parameters as needed; new parameters and the calculations for these parameters can be added to one of the existing result classes, and the software automatically adjusts the output files to accommodate the new variables. Adding a new parameter requires minimal extra code beyond that used to compute the result (further details can be found in the BRAVEHEART User Guide which is available on the BRAVEHEART GitHub).

### 2.14. VCG Signal and Annotation Quality Checks

BRAVEHEART was designed for batch processing of large numbers of ECGs with mininmal need for human oversight and without the need to manually review every ECG that is processed for signal quality, beat labeling, or removal of PVCs/outliers. The program can also take advantage of MATLAB’s parallel processing abilities which can significantly speed up processing of large ECG datasets. As ECG quality varies, especially when using clinical ECGs that were not collected specifically for research purposes, some lower quality ECG recordings are more likely to have errors in fiducial point annotation or median beat construction, usually due to significant artifact. In large studies it may not always be feasible to manually review every ECG that is processed, and we therefore designed ways of highlighting ECGs that are most likely to be problematic and require either exclusion from the dataset due to overall poor signal quality, automatic reannoation with other annotation parameters, or in very rare cases, manual ECG processing.

Further details of the quality checking methodology can be found in the “Quality Labeling Methods” section of the Supplemental Methods (Supplemental Section 1.7). Briefly, annotated median beats are automatically checked for possible processing errors or missannotation by checking a variety of metrics including the QT interval, QRS duration, T peak to QT ratio, T wave magnitude, average NCC between beats that make up the median beat, HR, number of beats included in the median beat construction, number of beats removed during PVC and outlier exclusion, presence of likely missing leads, and low neural network annotation confidence. If any of these values are outside of a nominal range (which can be easily adjusted via an external file as needed and based on the characteristics of the ECGs being processed), then the ECG is flagged for review. To streamline the review process, a separate file listing these flagged ECGs is generated, and figures from flagged ECGs can be automatically saved to a separate folder to facilitate review of flagged ECGs without needing to review the output of the entire dataset.

We also created a logistic regression based on a set of 481 sequentially processed ECGs/median beats which were manually labeled as “good” quality or “needs review” which can be used to predict the probability that the ECG being processed requires manual review (See Results Section 3.3, Supplemental Methods Section 1.7, Supplemental Results Section 2.1, Figure 10, and Supplemental Figure 4). ECGs that have a low probability of being “good” quality (based on an adjustable cutoff) can be flagged for review. This quality checking system is nominally designed to be specific, in that ECGs that have no flags are very unlikely to have any quality issues. Some ECGs that are flagged, however, may be perfectly fine after manual review or a minor adjustment. The thresholds for flagging ECGs can be adjusted by the user as needed if more specificity or more sensitivity is needed for a specific project (see Figure 10) and based on the types of ECGs being processed (e.g. are there frequent PVCs that will be removed or are all ECGs bradycardic), or the overall quality of the ECG recordings. This quality checking system can also be used to select ECGs with specific features (such as QRS duration *>* 150 ms or large amplitude T waves) from an unlabeled ECG dataset.

**Figure 10:**
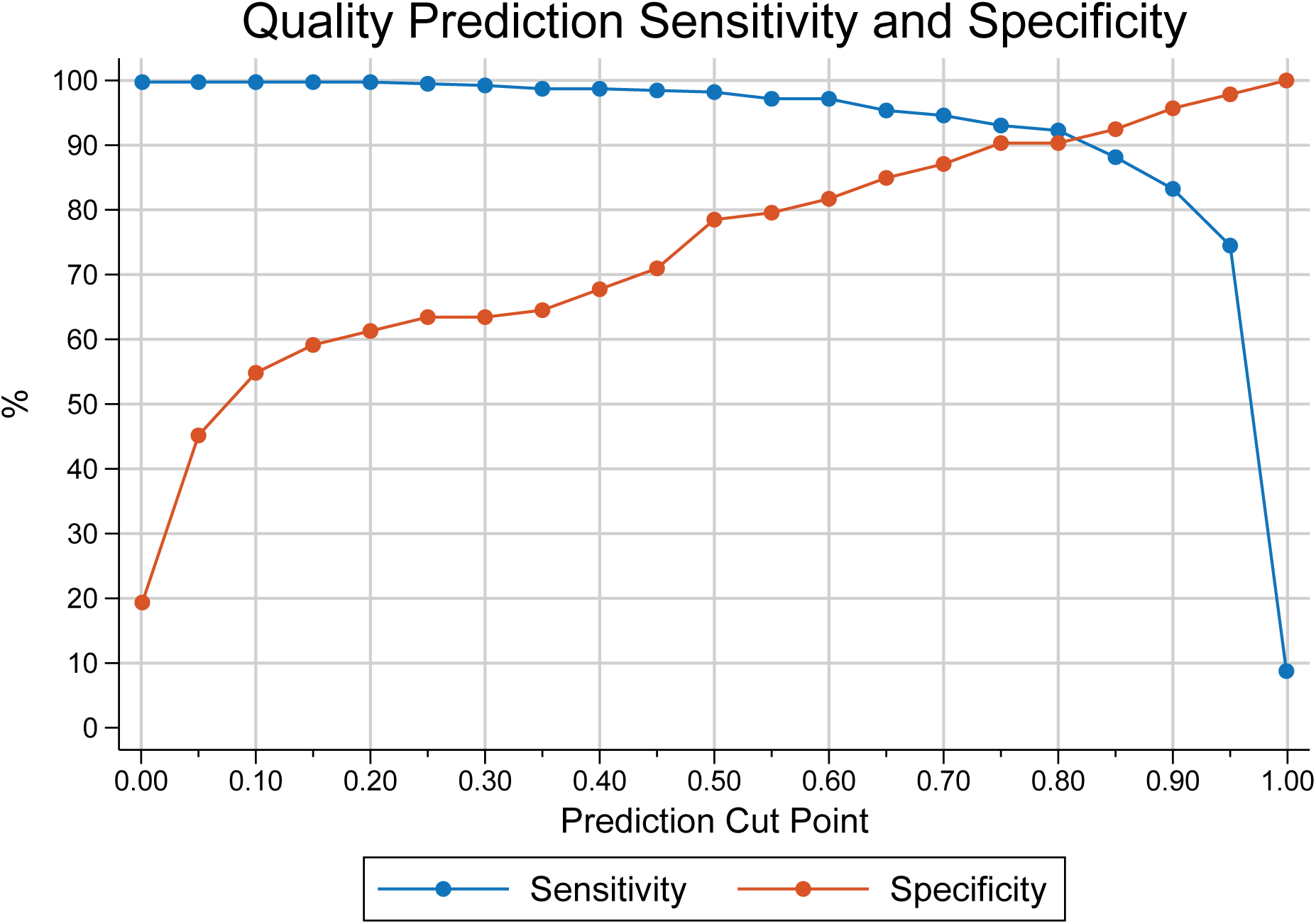
Sensitivity and specificity of the quality regression for predicting “good” quality ECGs. A cut point of 0.8 was set as nominal based on the relatively high and equal sensitivity and specificity at this point. No ECGs had a predicted probability of exactly 0 or exactly 1.00, so the point near “0.00” on the X-axis is for a cut point of 0.01, and the point near “1.00” on the X-axis is for a cut point of 0.99.

### 2.15. Graphical Display of VCG Median Beats and Export of Measurement Data

The BRAVEHEART software pipeline automatically produces a figure illustrating and summarizing the annotation process for each ECG/VCG, and example summary figures are shown in Figure 11 and Supplementary Figure 5. These summary figures can be quickly inspected to determine the number of beats included/excluded, why beats were excluded (PVC detection, outlier detection, manually removed, or missed), the accuracy and quality of median beat construction (via average NCC for all of the beats that made up the X, Y, and Z median beats), and the location of the median beat fiducial points. A detailed graphical user interface (GUI) that allows more specific adjustments and multiple additional visualizations is also available (see below).

**Figure 11:**
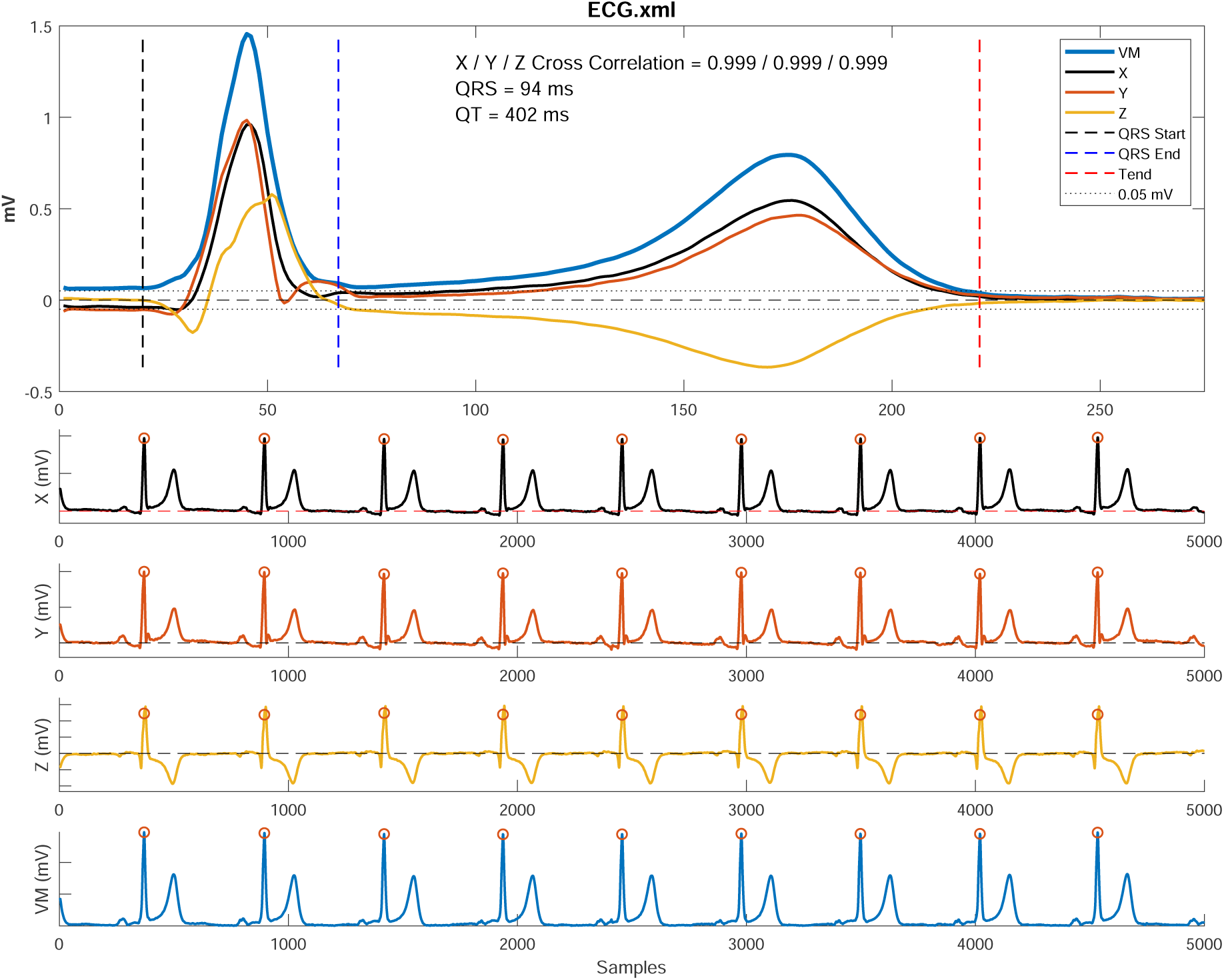
Example of summary figure output. The X, Y, Z, and VM median beats and fiducial point annotations are show in the top panel, and the full VCG is shown in the bottom half of the figure. Beats included in the median beat have an orange circle at their R peak. Beats excluded from the median beat (none in this example) would be annotated with the reason for exclusion (PVC or outlier). Cross correlation represents the average normalized cross correlation between all pairs of beats (in the X, Y, and Z leads) that make up the median beat and represents the quality of median beat construction with values very close to 1 indicating excellent beat alignment. **Abbreviations**: ECG - electrocardiogram, VCG - vectorcardiogram, VM - vector magnitude,

Measurement data for the processed ECG/VCG, which include the ECG filtering/processing settings used, are then exported to a file (.csv or .xlsx). ECGs processed in the same batch output their data to a single file for further analysis. The software can also save the ECG, VCG, and median beat signal data and ECG annotations to a separate file for additional processing/analysis outside of BRAVHEART.

The BRAVEHEART GUI (Figure 12) allows granular control of each step in ECG processing, including filtering, baseline correction, VCG construction, and beat removal, and can be quite helpful for troubleshooting ECGs which result in errors, or to better visualize each step in ECG/VCG processing. The GUI also allows visualization of different steps in ECG/VCG processing and additional figures, including standard 12-lead ECGs and VCGs, rotatable 3-dimensional VCG loops (Supplemental Figure 6), ECG/VCG lead morphology (Figure 9), and various figures describing the filtering and signal quality.

**Figure 12:**
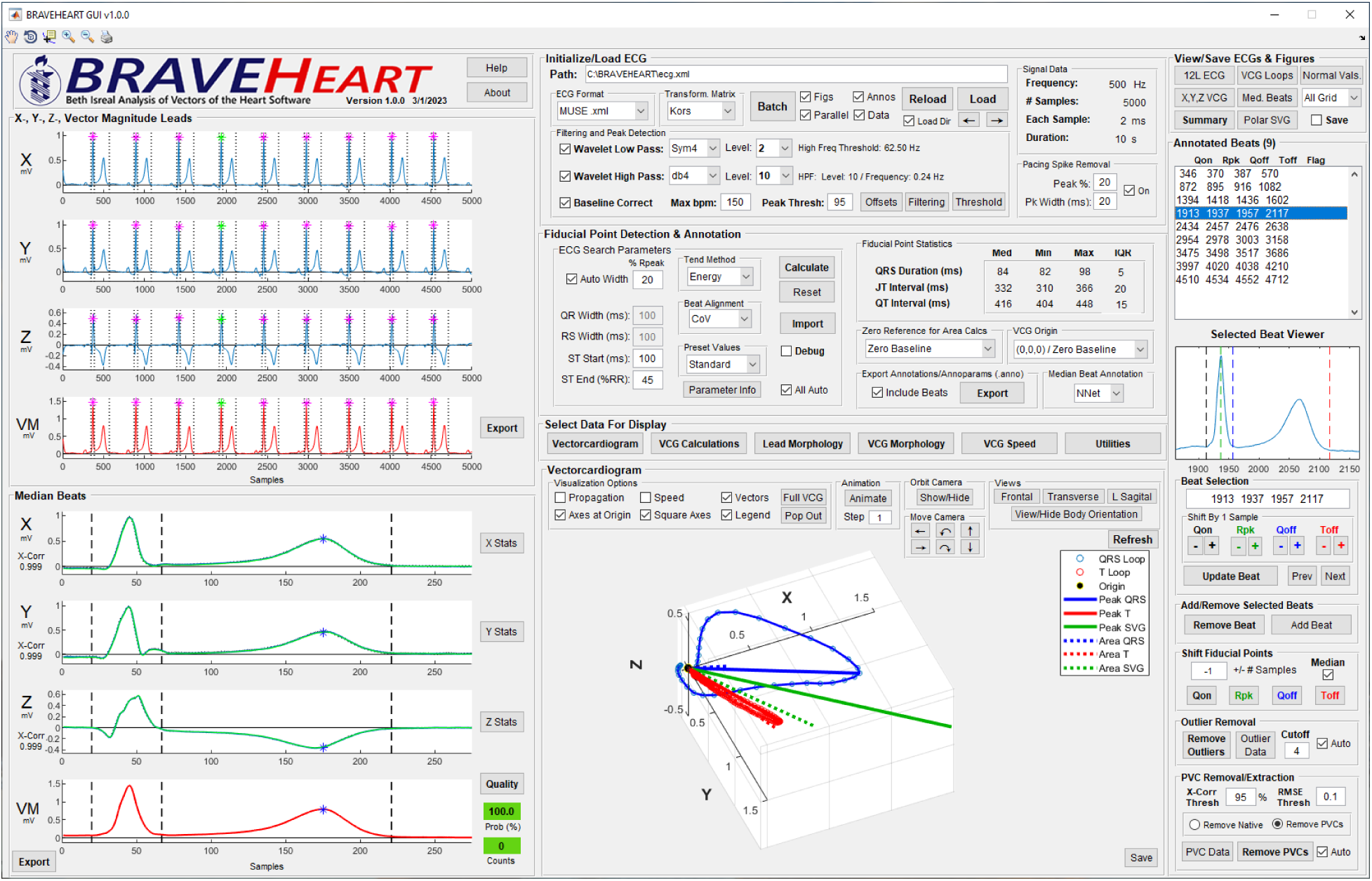
BRAVEHEART GUI. Further details on using the GUI can be found in the online User Guide.

### 2.16. Software Formats

BRAVEHEART is implemented in MATLAB and requires a version of at least R2022a. We have provided 2 packages that will suit different users. braveheart batch.m is a command line driven program that is designed for rapid, batch processing of ECGs with no graphical interface. Parameters are set in an external file, a directory is chosen for source ECG files, and the program processes the ECGs. The summary figure (Figure 11 and Supplimental Figure 5), ECG, VCG, and median beat signal data stored as .mat files, and annotation files (.anno) which contain the parameters used for ECG processing and individual beat fiducial point annotations for each processed ECG can be saved if specified by the user. ECGs that are not successfully processed by the completely automatic method and nominal settings can have their parameters manually adjusted, and individual beats can be removed/edited without the need for a GUI by editing an annotation (.anno) file that is placed in the same directory as the corresponding ECG. If the program notes the presence of an .anno file with the same name as the ECG being processed, the annotation parameters and fiducial point locations (if included) are read from the .anno file for that specific ECG. This allows the user control over ECGs that require non-nominal settings without requiring the use of the GUI.

We also provide a detailed GUI (braveheart gui.m) (Figure 12) that can perform both batch and individual ECG processing. The GUI is designed to give the user very fine control over each step of ECG processing including very granular control over beat annotation and removal. The GUI also allows additional visualizations of both ECG/VCG signal data and resulting measurements. A detailed overview of the GUI is available in the User Guide which is available on the BRAVEHEART GitHub repository. For users who do not have access to MATLAB, we have provided executable files for both the command line and GUI versions of BRAVEHEART for both Windows and Mac operating systems.

The complete BRAVEHEART MATLAB source code, executable files for Windows and Mac, and a detailed user guide, are available for download from GitHub http://github.com/BIVectors/BRAVEHEART under version 3 of the GPL license. BRAVEHEART uses the MATLAB signal processing, wavelet, and deep learning toolboxes. The parallel computing toolbox is optional to improve the speed of processing large batches of ECGs in parallel.

### 2.17. Statistical Analysis

All ECG processing and NN training were performed using MATLAB 2020b (Mathworks, Natick, MA, USA). Statistical analysis and data manipulation were performed using Stata 17 (StataCorp, College Station, TX, USA). The study was approved by the Institutional Review Board of Beth Israel Deaconess Medical Center.

## 3. Results

### 3.1. PVC Detection Performance

Supplemental Table 4 shows performance of the BRAVEHEART non-dominant QRST morphology (PVC) detector using various cutoffs of NCC and RMSE based on the summary performance of 1,000 replications of bootstrapping. In general, the best performance was found using a NCC of *≥* 95%, and a RMSE = 0.1, which is why these values were chosen for nominal settings. Performance was also excellent when NCC was disabled, and decisions on similarity were based on RMSE alone. With NCC = 95% and RMSE = 0.1, sensitivity was 98.2%, specificity was 99.5%, and F1 was 97.7% for PVC detection. Although PVC detection performance was excellent using a RMSE cutoff of 0.1 with or without use of NCC in certain cases it can be advantageous to utilize both NCC and RMSE, as a beat with excellent matches by both NCC and RMSE may be a better template beat than if only matched by RMSE (See Supplemental Figure 12).

### 3.2. Neural Network Performance

Supplemental Table 5 shows the features/diagnoses of ECGs used for NN training and testing during NN development. There were overall no significant differences in ECG features between the training and testing datasets with the exception of a borderline lower rate of left bundle branch block ECGs in the testing dataset (3.2 vs 7.1 %, *p*=0.046). Table 1, Figure 13 and Supplemental Figures 7, 9, and 8 show the accuracy of NN fiducial point annotations when compared to ground truth human annotations for the 189 ECGs set aside during NN training. There was excellent agreement between NN predicted and ground truth fiducial point locations (*Q*_on_, *Q*_off_, and *T*_off_) and ECG intervals (QRS duration and QT interval) with mean differences *< ±* 2 ms for all fidual points/intervals, and all measurement error distributions were well within established acceptable limits for automated ECG annotation [27, 28, 29, 30]. Microaveraged F1 (*µ*F1 - See Supplemental Statistical Methods in Supplement Section 1.8) was 97.95%. The NN did not miss locating any fiducial points, and only 1 ECG was found to have more an extra fiducial point (1 *T*_off_) detected which was appropriately dealt with using logic that accounts for the order of fiducial points. Supplemental Figure 7 shows intraclass correlation coefficients for all fiducial points and intervals were very close to 1.0. Supplemental Figure 8 shows the percent of annotations that were within different intervals of accuracy; more than 99% of *Q*_on_ and *Q*_off_ annotations and more than 98% of *T*_off_ annotations were within 20 ms of ground truth. Bland-Altman plots for NN performance are available in Supplemental Figure 9.

**Figure 13:**
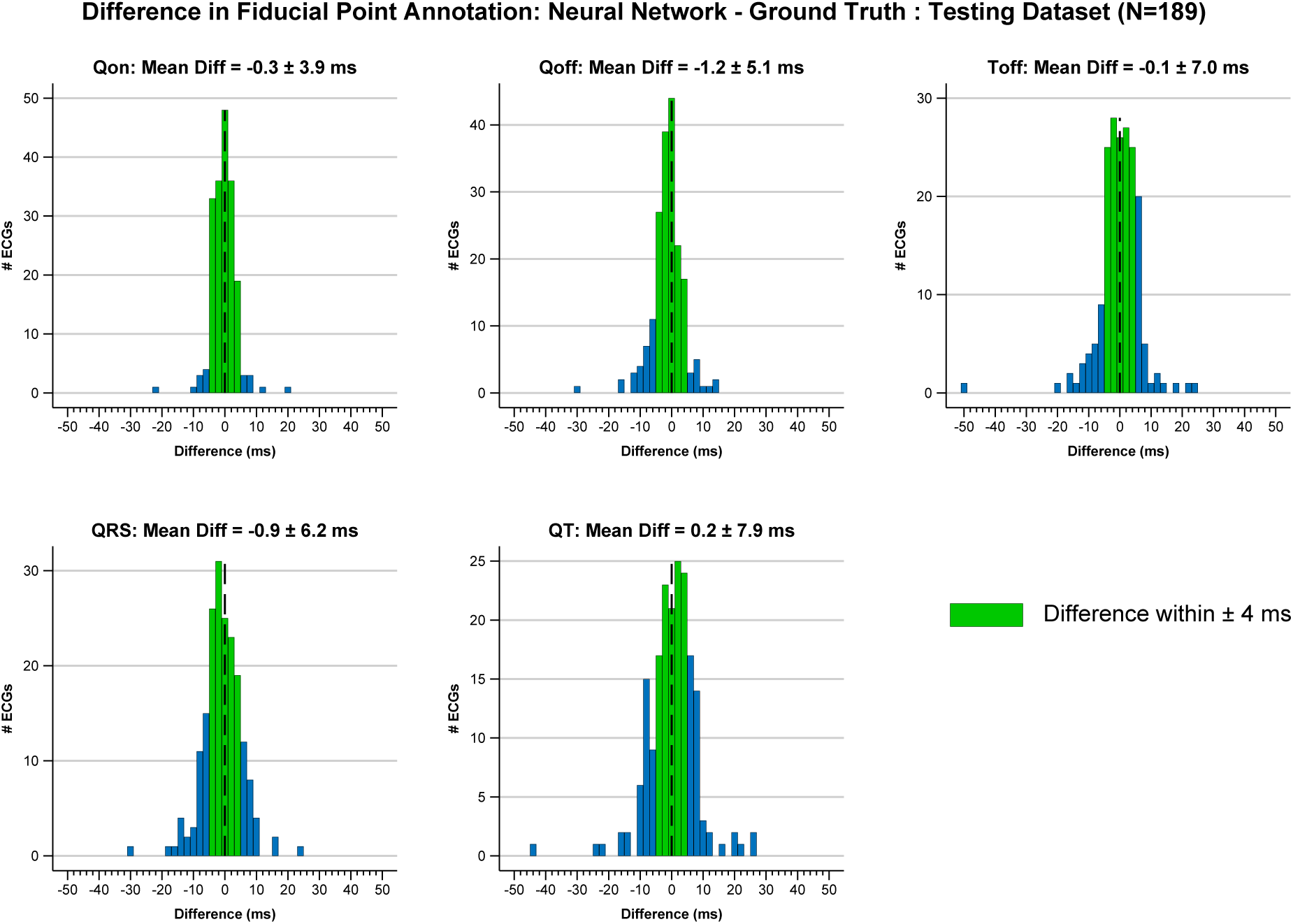
Mean and standard deviation and histograms for results of NN testing for the initial 189 ECG testing dataset. Bins where the difference between NN predictions and ground truth fiducial points were *≤ ±* 4 ms are shown in green.

**Table 1:**
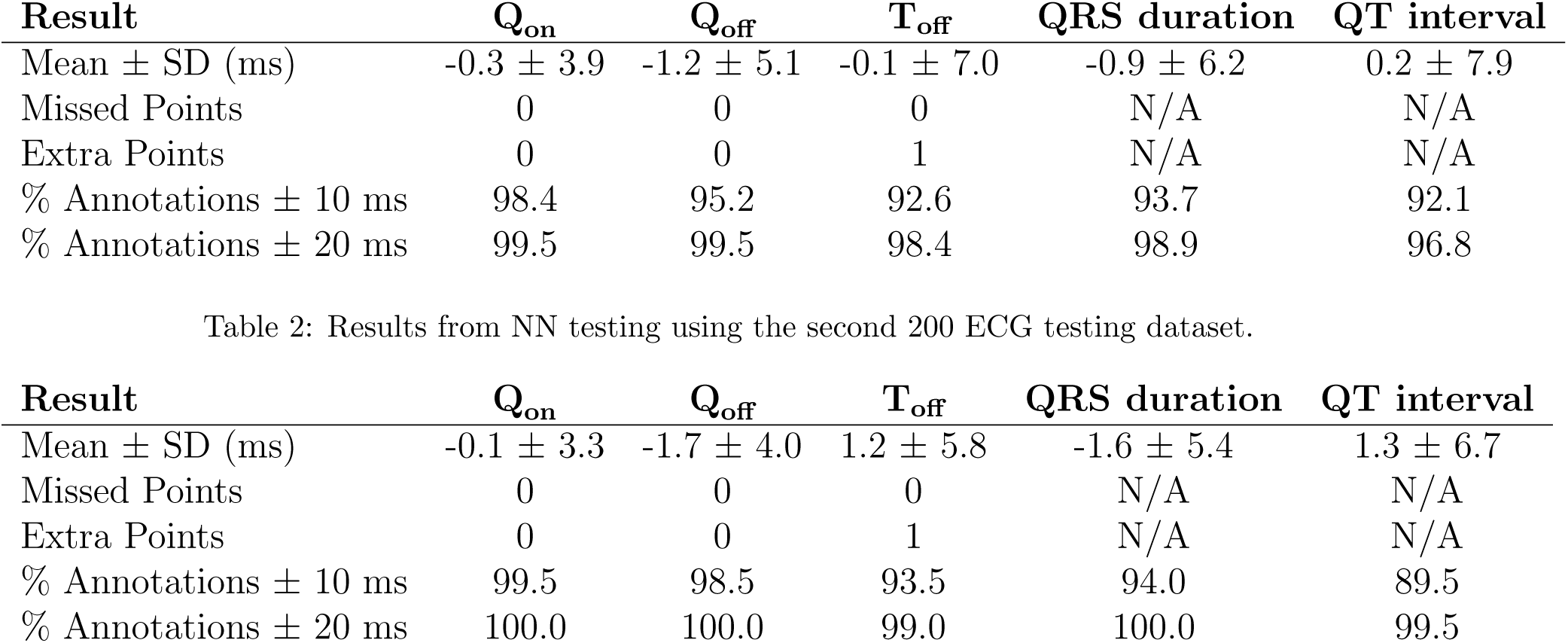
Results from NN testing using the initial 189 ECG testing dataset.

Supplemental Table 6 shows the features/diagnoses of ECGs included in the second, independent testing dataset of 200 sequentially obtained ECGs. Table 2 shows that the NN performed similarly on this second dataset, with mean differences *< ±* 2 ms for all fiducial points/intervals. More than 99% of all fiducial points and intervals were within 20 ms of ground truth. *µ*F1 was 98.03%. The NN did not miss locating any fiducial points, and only 1 ECG was found to have an extra fiducial point (1 *T*_off_) detected which was appropriately dealt with using logic that accounts for the order of fiducial points. Figures summarizing the performance of the second dataset are shown in Supplemental Figures 10 and 11.

### 3.3. Quality Labeling Results

Figure 10 and Supplemental Figure 4 show the results for identifying an ECG/median beat as “good quality” or “needs review” using the previously discussed logistic regression (See Supplemental Sections 1.7 and 2.1). The cutoff used to distinguish “good quality” and “needs review” ECGs can be adjusted as needed for a specific project to increase/decrease sensitivity or specificity. In general, a cutoff of *∼*80% tends to have a good mix of sensitivity and specificity, with both values *>*90%. Cutoff values *>*90% have higher specificity at the expense of lower sensitivity, and cutoff values *<*70% have higher sensitivity at the expense of lower specificity. The ROC curve for the quality regression is shown in Supplemental Figure 4 and had an area under the ROC of 0.973 indicating excellent agreement between prediction and manual labeling. Further details can be found in the Supplemental Methods/Results (Supplemental Sections 1.7 and 2.1).

## 4. Discussion

We present BRAVEHEART, an easy to use and easily customizable research software package for ECG and VCG analysis. BRAVEHEART satisfies an important need in the development of ECG/VCG research by providing open-source software that performs all signal processing and analyses itself without the need for separate programs to process or annotate ECG signals. The software accepts a wide variety of ECG formats, and is easily customizable with minimal extra programming. We also have provided executable versions of the software for those without access to MATLAB. Extensive documentation is available in a user guide to help users familiarize themselves with the software. Although other ECG/VCG processing software does exist [8, 9, 12, 13, 10, 11], these other software packages either are not publically available, not open-source, or provide partial functionality without allowing complete processing from ECG to VCG and final measurements. We are aware of the importance of being able to modify the software and add additional measurements based on specific research needs, and have taken care to make it easy to add new measurements with minimal effort and coding (see the online user guide).

Pre-release versions of BRAVHEART have been used in research investigating associations between VCG parameters (primarily the SVG) and drug induced ventricular arrhythmias [31], antiarrhythmic drug administration [20], post pulmonary embolism risk stratification [32], and chemotherapy-induced cardiotoxicity [33], and the current release version of BRAVEHEART is being used for ECG/VCG analysis in multiple ongoing studies. We hope that members of the community will help improve BRAVEHEART over time by suggesting or adding new or improved features.

## 5. Limitations/Future Directions

BRAVEHEART currently does not support P wave annotation. We plan to add this as a feature in the near future.

## 6. Conclusion

BRAVEHEART is an open-source, easily customizable ECG/VCG analysis software package that reproducibly processes digitial 12-lead ECG files without the need for external software libraries or signal pre-processing. Source code, executables, and a detailed user guide are available at http://github.com/BIVectors/BRAVEHEART where it is distributed under the General Public License (GPL) version 3.

## Supporting information

Supplemental Methods and Results

## Data Availability

Source code for this work is available online at https://github.com/BIVectors/BRAVEHEART

