## Supplemental Methods and Results for "BRAVEHEART: Open-source software for automated electrocardiographic and vectorcardiographic analysis"

Hans Friedrich Stabenau, MD, PhD and Jonathan W. Waks, MD
Harvard-Thorndike Electrophysiology Institute
Beth Israel Deaconess Medical Center
Harvard Medical School, Boston, MA

### 1. Supplemental Methods

#### 1.1. Heuristic Annotation Details:

Once the QRS search window is established, for each QRS complex, the start ($Q_{\text{on}}$) and end ($Q_{\text{off}}$) are determined with the following method. First, the algorithm searches to determine if there is an additional peak in the VM lead before the dominant R wave peak within a short distance (i.e. a Q wave – all peaks are positive in the VM lead). Two methods are then used to determine the start of the peak. Method A finds the maximum slope of the R peak and walks forward/back until it finds a point where the absolute value of the derivative is 2% of the maximum slope (or the smallest value found if not < 2%). Method B looks for the first local minimum and labels this as the fiducial point. The point closest to dominant VM R peak from methods A and B is chosen as the fiducial point.

After $Q_{\text{on}}$ and $Q_{\text{off}}$ are located, a blanking window (nominally 100 ms) is set after the location of $Q_{\text{off}}$ during which the T wave end ($T_{\text{off}}$) cannot be detected. At the end of this blanking window, the $T_{\text{off}}$ search window starts and is extended forward nominally by 45% of the mean RR interval. The $T_{\text{off}}$ search window is set as a % of the mean RR interval rather than an absolute value in ms to minimize the need to change parameter values as the QT interval is related to the RR interval/HR. The location of $T_{\text{off}}$ is then detected by the chosen method as noted in the main manuscript. Fiducial point search windows are illustrated in Figure 4 in the main manuscript.

#### 1.2. PVC Detection Algorithm Details:

First, The $n$ segmented QRST complexes that are used to create the median beat are trimmed around their center of voltage (CoV - see Equation 3 in the main manuscript) to the shortest QR and RT intervals from all $n$ beats, so that all segmented beats are the same length. This is required to perform cross correlation, and since the QRS complex is the most important feature to perform QRST beat comparison, trimming short lengths of signal from the start or end of the beat is not detrimental. Trimming the beats also reduces the influence of long isoelectric segments where artifact or signal offsets can cause poor matching.

Figure 6, and Supplemental Figures 2 and 3 provide a visual overview and explanation of the PVC detection algorithm. Each of the $n$ beats is compared to itself and the remaining $n-1$ beats using NCC and RMSE normalized to the max-min of the signals being compared. This results in a $n \times n$ matrix ($C$) of NCC values and a second $n \times n$ matrix ($R$) of RMSE values. NCC values above a set threshold (nominally 95%) and RMSE values below a set threshold (nominally 0.1) are assigned a value of 1, while beat-to-beat comparisons that do not meet these criteria are assigned a value of 0. After thresholding, matrices $C$ and $R$ are therefore both $n \times n$ matrices with values of either 0 or 1; beat pairs that have a value of 1 are considered to have similar morphologies based on meeting either NCC or RMSE criteria, while beat pairs that have a value of 0 are considered to have dissimilar morphologies. The matrices $C$ and $R$ are then added together to form a new $n \times n$ "template matrix" ($T$) which has values of 0, 1, or 2 based on whether the beat pair was considered similar by neither NCC or RMSE, either NCC or RMSE, or both NCC and RMSE, respectively.

The $n$ rows of template matrix $T$ are summed, so that there is now a $n \times 1$ matrix where beats that are correlated with more of the remaining $n-1$ beats have higher values compared to beats that are correlated with fewer other beats. The first maximum value of this $n \times 1$ matrix is set as the "reference" beat (beat $n_{\text{ref}}$), and the corresponding row of matrix $T$ is saved as a corresponding $1 \times n$ "dominant morphology" matrix $D$ where the values of 0, 1, or 2 indicate the strength of similarity of the reference beat $n_{\text{ref}}$ to the remaining $n-1$ beats. Another thresholding procedure sets values of matrix $D$ which are $\geq 1$ equal to 1, and does not change values of 0. Matrix $D$ therefore is now a $1 \times n$ matrix with values $D_i$ equal to 0 if if the reference beat had no correlation by either NCC or RMSE with the $i$th beat, and $D_i$ equal to 1 if the reference beat had correlation by NCC and/or RMSE with the $i$th beat.

The software starts by assuming that the reference beat, $n_{\text{ref}}$, by virtue of it being correlated with the largest number of other beats, represents the "dominant", non-PVC QRST morphology rather than PVC/paced/aberrant QRST morphology. If the number of "dominant" beats (the number of 1s in $D$) is $> n/2 + 1$, the software assumes that the reference beat QRST morphology is truly the "dominant"/"normal" QRST morphology for the ECG as it is present in the majority of beats. Beats with a dominant matrix ($D_i$) value of 1 are therefore assigned to be dominant beats, while beats with a $D_i$ value of 0 are assigned to be PVCs.

If $\leq n/2 + 1$ beats are "dominant" based on matrix $D$, there is a possibility that the reference beat is actually a PVC because there may be similar numbers of normal beats and PVCs (such as occurs with ventricular bigeminy), and/or the PVCs may be more similar with each other than the normal beats are with each other due to noise, differences in alignment, or other factors. In this case, to determine which template is truly the "dominant" QRST morphology, the software compares the QRS durations and QT intervals for the best matches of the dominant and non-dominant beat morphologies. Beat $n_{\text{ref}}$ is chosen as the template for the "dominant" QRST template, and a similar selection procedure is performed on "PVC" beats to find the beat that has the highest correlation with all other PVC beats. If the QRS interval of the "dominant" template beat is $< 90\%$ of the QRS interval of the "PVC" template beat, the reference beat is defined as "dominant" and values of 0 in matrix $D$ are identified as PVCs. If the QRS interval of the "dominant" template beat is not $< 90\%$ of the QRS interval of the "PVC" template beat, the software does the same comparison with the QT interval, and if the QT interval of the "dominant" template beat is $< 90\%$ of the QT interval of the "PVC" template beat the reference beat is defined as "normal" and values of 0 in matrix $D$ are identified as PVCs. Otherwise the software decides that it initially selected the "dominant" QRST morphology incorrectly, and values of 1 in matrix $D$ are identified as PVCs.

#### 1.3. Outlier Detection Using Modified Z-Scores:

For each beat, the QR and RT intervals are computed. A modified Z-score ($Z_i$) for each of these quantities is then computed based on the median absolute deviation (MAD) or mean absolute deviation (MeAD) [1]:

$$\begin{aligned} \text{MAD} &= \text{median}(|x_i - \tilde{x}|) \\ \text{MeAD} &= \text{mean}(|x_i - \bar{x}|) \end{aligned} \tag{1}$$

where $x_i$ is the value for the $i$th beat, $\tilde{x}$ is the median value for all $i$ beats, and $\bar{x}$ is the mean value for all $i$ beats.

For each QR or RT interval:

$$\begin{aligned} Z_i &= 0.6745 * (x_i - \tilde{x})/\text{MAD} \qquad \text{if MAD} \neq 0 \\ Z_i &= 0.7979 * (x_i - \tilde{x})/\text{MeAD} \qquad \text{if MAD} = 0 \end{aligned} \tag{2}$$

If $Z_i$ is $> 3.5$ nominally [1] (this cutoff point can be adjusted) the $i$th beat is marked as an outlier and can be automatically removed.

#### 1.4. Neural Network Training Data Source/Preparation:

To train the vector magnitude (VM) median beat annotator neural network (NN), we searched the Beth Israel Deaconess Medical Center ECG database (MUSE, General Electric, Boston, MA) and obtained 1,262 sequential, 10-second 12-lead ECGs sampled at 500 Hz with a 4.88 $\mu$V/unit resolution which were of diagnostic quality. ECGs with extreme noise, missing leads, or other technical issues were excluded. Using a pre-release version of BRAVEHEART, 12-lead ECGs were first filtered with both low- (62.5 Hz) and high-pass (0.24 Hz) wavelet filters using the Symlets 4 and Daubechies 4 wavelets, respectively, to remove noise and baseline wander. ECGs were transformed into orthogonal X, Y, Z leads using the Kors transformation matrix [2]. The median VM lead was constructed from the median X, Y, and Z leads as described in the main manuscript methods. Determination of ECG fiducial points is most unambiguously performed on the VM lead [3] and use of the VM lead allows assessment of “global” fiducial points and intervals on a single lead [4]. Use of the VM lead also simplifies computation needed for fiducial point detection because the VM lead is always positive, and VM lead morphology is simpler/more conserved than the variety of possible signal morphologies seen in the 12 standard ECG leads.

Median VM beats were constructed by aligning individual beats based on their center of voltage (CoV) as described in the main manuscript. QRS onset ($Q_{\text{on}}$), QRS offset ($Q_{\text{off}}$), and T wave offset ($T_{\text{off}}$) were then manually annotated by an experienced, board certified cardiac electrophysiologist (JWW) using electronic calipers, and the annotated median beat signals and annotations were exported for later use. We included normal and abnormal ECGs, including paced ECGs. For paced QRS complexes, $Q_{\text{on}}$ was defined as occurring just after the completion of the high-frequency pacing spike. P waves were not annotated.

Each sample in the VM waveform was associated with a categorical signal containing one of three labels; "QRS complex", "T wave," or "other" based on the annotated fiducial points. "QRS complex" was defined as the signal between $Q_{\text{on}}$ and $Q_{\text{off}}$, "T wave" was defined as the signal between $Q_{\text{off}} + 1$ sample and $T_{\text{off}}$, and all other parts of the VM waveform were defined as "other" (see Figure 8 in the main manuscript).

#### 1.5. Neural Network Architecture and Training:

We trained a bidirectional Long-Short Term Memory (biLSTM) NN [5] for sequence-to-sequence classification using Matlab version 2020b. Details of the chosen NN architecture and its performance have been previously reported [6]. The 1,262 annotated median VM beats from 1,262 unique patients were split into training (75%, n=947), validation (10%, n=126), and testing (15%, n=189) datasets. The training dataset beats were then augmented using previously described and validated median beat ECG augmentation software (ECGAug) [6] by recombining different QRS and T wave segments with further physiologically appropriate distortion while maintaining the locations of manually annotated fiducial points. Augmentation resulted in a final training dataset of 8,479 annotated VM median beats. Prior to training, the VM median beat signals were further processed by removing frequencies outside of 0.5-40 Hz (the power spectrum of the QRST complex)[7] using the Fourier synchrosqueezed transform with a Kaiser-Bessel window of length 128 and $\beta$=0.5, and then standardized by subtracting the mean and diving by the standard deviation.

For NN training, the 20 real and imaginary parts of the transformed signal were used as input into the biLSTM. The input layer was fed into a biLSTM layer with 80 neurons which connected to a fully-connected layer with 3 outputs and then a softmax activation function to assign probabilities for the 3 possible labels ("QRS complex", "T wave", or "other") at each sample of the VM median beat signal. Hyperparameters were manually tuned to find optimal NN performance based on prior experiments [6]. The final/best NN was trained with a batch size of 32 for 5,000 iterations using the Adam optimizer [8]. The initial learning rate was 0.003 with a 80% decrease in learning rate every 400 iterations. We empirically found that more layers and/or more neurons resulted in over-fitting of the data, while fewer neurons resulted in poorer annotation performance. Forcing training to proceed above 5,000 iterations tended to result in over-fitting [6]. Training curves for the final NN are shown in Supplemental Figure 13.

Just as human fiducial point annotations were used to create a categorical signal for NN training (Figure 8A in the main manuscript), the NN output probabilities of each of the 3 labels ("QRS complex", "T wave", or "other") were then used to assign NN predicted fiducial points based on the locations of transition between segments of the QRST complex (Figure 8B in the main manuscript). Logic was used to convert these probabilities into fiducial point locations. In general, $Q_{\text{on}}$ was assigned as the sample where there was a transition in the predicted categorical signal from "other" to "QRS complex", $Q_{\text{off}}$ was assigned as the sample where there was a transition in the predicted categorical signal from "QRS complex" to "T wave", and $T_{\text{off}}$ was assigned as the sample where the predicted categorical signal transitioned from "T Wave" to "other". For signals that are not sampled at 500 Hz, the signal is down/upsampled to 500 Hz before being passed into the NN, and the calculated fiducial points are then adjusted based on the original frequency.

In rare cases where there were multiple points where the definition for a fiducial points was satisfied (usually ECGs with significant artifact or very low T wave amplitude), an algorithm which takes the relative locations of $Q_{\text{on}}$, $Q_{\text{off}}$, and $T_{\text{off}}$ was used to choose the optimal location for the fiducial point; for example, the algorithm does not allow assignment of fiducial points out of order, and will not assign a $T_{\text{off}}$ before a $Q_{\text{off}}$. These ECGs are flagged for review as part of BRAVEHEART post processing quality checking (see Methods of the main manuscript for details).

#### 1.6. Neural Network Performance Assessment:

Since we were primarily interested in fiducial points which occur at transitions between labels in the QRST complex, overall classification accuracy for the entire VM signal was not felt to be a good measure of NN performance. The majority of the VM median beat signal would always be annotated correctly regardless of NN performance, but the predicted fiducial points could differ significantly from ground truth even with high overall classification accuracy for the entire signal; for example, if a median beat signal was 500 samples, $Q_{\text{on}}$ could be off by 10 samples (20 ms at 500 Hz), which would indicate overall poor performance, but the entire signal could still be annotated with 99% accuracy.

NN performance was therefore assessed by comparing the predicted location of $Q_{\text{on}}$, $Q_{\text{off}}$, and $T_{\text{off}}$ and the clinically relevant intervals (QRS duration $[Q_{\text{off}} - Q_{\text{on}}]$ and QT interval $[T_{\text{off}} - Q_{\text{on}}]$) to the ground truth expert manual annotations/intervals in the testing dataset. Unlike many other classification problems where there is an unambiguous ground truth for use in NN training and testing, fiducial point annotation is associated with inherent uncertainly, and even trained experts can sometimes disagree as to the true location of a specific fiducial point (especially the T wave end) [9]. Thresholds for adequate performance were based on consensus statements for electronic ECG annotation [10, 11, 12, 13].

The mean and standard deviation of the difference between the NN predicted and ground truth fiducial point locations and intervals were calculated. Missing transitions were defined as when a $Q_{\text{on}}$, $Q_{\text{off}}$, or $T_{\text{off}}$ fiducial point was not found in a median VM QRST signal, and extra transitions were defined as when multiple potential $Q_{\text{on}}$, $Q_{\text{off}}$, or $T_{\text{off}}$ fiducial points were present in a single median VM QRST signal. As this was a multi-class labeling problem with imbalanced classes, the precision (positive predictive value), recall (specificity), and F1 (the harmonic mean of precision and recall) are all equal, and only micro-averaged F1 was therefore reported (See Supplemental Statistical Methods below).

We have previously described detailed training results using the current biLSTM NN architecture [6], and given that BRAVEHEART relies heavily on using the specific weights and biases which are included in the software, for the purpose of the software release we validated NN performance on a second, independent testing dataset using 200 sequential ECGs obtained a single day which were free of significant artifact or missing leads. NN predicted fiducial points and intervals were compared to ground truth annotations for this dataset in the same way as noted above for the original testing dataset.

#### 1.7. Quality Labeling Methods:

A set of 481 sequential ECGs obtained on a single day and which were not used for any other parts of BRAVEHEART development were processed by BRAVEHEART using nominal settings and without regard to signal quality. Processed ECGs/VM median beats were manually reviewed and labeled as “good quality” or “needs review”. Multivariable logit regression was performed, initially including the minimum NCC for the X, Y, and Z leads (NCC), median VM lead voltage in the 30 ms after the end of the T wave (baseline), maximum VM lead T wave voltage (TMag), VM lead T peak-T end ratio (TPTE), number of removed beats (either via PVC or outlier detection), and measures of high-frequency and low-frequency noise. After adjustment, only NCC, baseline, TMag, and TPTE were independently associated with the quality of the final processed ECG. The final logit coefficients were used to calculate predicted probabilities (range 0–1) of “good quality” ECGs after BRAVEHEART processing, and receiver-operating curves (ROC) and sensntivity and specificity were calculated at various cut points.

#### 1.8. Supplemental Statistical Methods:

Microaveraged ($\mu$) Statistics help adjust for the imbalances present between different classes in the VM median beat annotation data:

$$\mu\text{Precision} = \frac{\sum_{k=1}^{3} \text{TP}_k}{\sum_{k=1}^{3} \text{TP}_k + \text{FP}_k} \tag{3}$$

$$\mu\text{Recall} = \frac{\sum_{k=1}^{3} \text{TP}_k}{\sum_{k=1}^{3} \text{TP}_k + \text{FN}_k} \tag{4}$$

$$\mu\text{F1} = 2 \times \frac{\mu\text{Precision} \times \mu\text{Recall}}{\mu\text{Precision} + \mu\text{Recall}} \quad (5)$$

where TP are true positives, FP are false positives, and FN are false negatives, and for $k$ = the 3 different categorical labels are "QRS Complex", "T Wave", or "other".

### 2. Supplemental Results

#### 2.1. Quality Labeling Results:

The final logit model was given as:

$$p = -22.56 + (18.21 * \text{NCC}) + (-87.59 * \text{baseline}) + (9.06 * \text{TMag}) + (11.19 * \text{TPTE}) \quad (6)$$

and therefore the probability of a specific processed ECG being "good" is given as:

$$P(\text{good}) = e^p/(1 + e^p) \quad (7)$$

The ROC curve for the quality regression is shown in Supplemental Figure 4 and had an area under the ROC of 0.973 indicating excellent agreement between prediction and manual labeling. Sensitivity and specificity at various cut points of $P$(good) are shown in Figure 10 in the main manuscript. As can be seen, at around a cut point of 0.8 there was a good combination of sensitivity and specificity, with both values $> 90\%$. Cut points $> 0.9$ tended to have higher specificity and lower sensitivity, while values $< 0.7$ tended to have lower specificity and higher sensitivity. Cut points can be adjusted in BRAVEHEART based on the dataset being analyzed and the relative sensitivity or specificity that is desired in the dataset being analyzed, which will depend on the overall quality of the ECG tracings being analyzed.

### 3. Equations

**Vector Magnitude:**

Vector magnitude (VM) is defined as the Euclidean norm of the VCG:

$$\mathrm{VM} = \sqrt{X^2 + Y^2 + Z^2} \tag{8}$$

**Area Vectors:**

Area vectors are obtained by taking the area under the relevant segment of the median VCG ($\mathbf{V}(t) = [X(t), Y(t), Z(t)]$) QRST complex using the trapezoidal rule:

$$\mathbf{QRS}_{\text{area}} = \int_{Q_{\text{on}}}^{Q_{\text{off}}} \mathbf{V}(t)\, dt = \left[ \int_{Q_{\text{on}}}^{Q_{\text{off}}} X(t)\, dt, \int_{Q_{\text{on}}}^{Q_{\text{off}}} Y(t)\, dt, \int_{Q_{\text{on}}}^{Q_{\text{off}}} Z(t)\, dt \right] \tag{9}$$

$$\mathbf{T}_{\text{area}} = \int_{Q_{\text{off}}}^{T_{\text{off}}} \mathbf{V}(t)\, dt = \left[ \int_{Q_{\text{off}}}^{T_{\text{off}}} X(t)\, dt, \int_{Q_{\text{off}}}^{T_{\text{off}}} Y(t)\, dt, \int_{Q_{\text{off}}}^{T_{\text{off}}} Z(t)\, dt \right] \tag{10}$$

**Peak Vectors:**

Peak vectors are obtained by taking the value of relevant segment of $\mathbf{V}(t)$ at the time point which is of maximum distance from the origin:

$$\mathbf{QRS}_{\text{peak}} = [X(t_{Qmax}), Y(t_{Qmax}), Z(t_{Qmax})] \tag{11}$$

$$\mathbf{T}_{\text{peak}} = [X(t_{Tmax}), Y(t_{Tmax}), Z(t_{Tmax})] \tag{12}$$

where $t_{Qmax}$ is the time of max distance of the QRS loop from the origin, and $t_{Tmax}$ is the time of max distance of the T loop from the origin. These times correspond to the maximum values of the QRS complex and T wave in the VM lead, respectively.

**Spatial Ventricular Gradient:**

The Spatial Ventricular Gradient (SVG) is the vector created by the QRST integrals in $X, Y$, and $Z$:

$$\mathbf{SVG} = \mathbf{QRS}_{\text{area}} + \mathbf{T}_{\text{area}} = \int_{Q_{\text{on}}}^{T_{\text{off}}} \mathbf{V}(t)dt = \left[ \int_{Q_{\text{on}}}^{T_{\text{off}}} X(t)\, dt, \int_{Q_{\text{on}}}^{T_{\text{off}}} Y(t)\, dt, \int_{Q_{\text{on}}}^{T_{\text{off}}} Z(t)\, dt \right] \tag{13}$$

**Azimuth Angle:**

Azimuth is defined as the angle in the transverse (XZ) plane with negative angles pointing anterior, and positive angles pointing posterior (see Supplemental Figure 14). Azimuth can take values from 0 to ±180 degrees, with 0 degrees pointing to towards the left, and ±180 degrees pointing towards the right.

$$\text{azimuth} = \arctan\left(\frac{Z}{X}\right) \tag{14}$$

**Elevation Angle:**

Elevation is defined as the angle in the frontal (XY) plane (See Supplemental Figure 14). Values range from 0 to 180 degrees, with 0 degrees pointing towards the feet and 180 degrees pointing towards the head.

$$\text{elevation} = \arccos\left(\frac{Y}{\text{VM}}\right) \tag{15}$$

**Absolute Integrals:**

The sum absolute integral (SAI) is defined as the area under the absolute value of the QRST complex:

$$\text{SAI}_i = \int_{Q_{\text{on}}}^{T\text{off}} |V_i(t)|\, dt \qquad \text{for } i = X, Y, Z, \text{ or VM} \tag{16}$$

SAI QRST is defined as:

$$\text{SAI QRST} = \text{SAI}_x + \text{SAI}_y + \text{SAI}_z = \int_{Q_{\text{on}}}^{T\text{off}} |X(t)|\, dt + \int_{Q_{\text{on}}}^{T\text{off}} |Y(t)|\, dt + \int_{Q_{\text{on}}}^{T\text{off}} |Z(t)|\, dt \tag{17}$$

**QRST Angles:**

The spatial QRST angle is the 3-dimensional angle between QRS and T vectors:

$$\text{QRST Angle} = \arccos\left(\frac{\mathbf{QRS} \cdot \mathbf{T}}{|\mathbf{QRS}|\,|\mathbf{T}|}\right) \tag{18}$$

where the peak QRST angle uses $\mathbf{QRS}_{\text{peak}}$ and $\mathbf{T}_{\text{peak}}$, and the area QRST angle uses $\mathbf{QRS}_{\text{area}}$ and $\mathbf{T}_{\text{area}}$.

**Total Cosine R to T (TCRT):**

TCRT was calculated as previously described using singular value decomposition [14].

**VCG Loop Length:**

Loop length is calculated as the sum of distances covered as the VCG loop increments by each sample For example, QRS loop length is calculated as:

$$\sum_{i=Q_{\text{on}}}^{Q_{\text{off}-1}} \sqrt{(X_{i+1} - X_i)^2 + (Y_{i+1} - Y_i)^2 + (Z_{i+1} - Z_i)^2} \tag{19}$$

and T loop length is calculated as:

$$\sum_{i=Q_{\text{off}}}^{T_{\text{off}-1}} \sqrt{(X_{i+1} - X_i)^2 + (Y_{i+1} - Y_i)^2 + (Z_{i+1} - Z_i)^2} \tag{20}$$

**VCG Loop Speed:**

The instantaneous speed of the QRS or T loops ($v_i$) in mV/ms for a VCG with frequency $f$ is calculated as the distance traveled in a sample of time:

$$v_i = \sqrt{(X_{i+1} - X_i)^2 + (Y_{i+1} - Y_i)^2 + (Z_{i+1} - Z_i)^2} \,/\, \Delta t \qquad \text{where } \Delta t = 1000/f \tag{21}$$

**Left Ventricular Hypertrophy (LVH):**

LVH metrics are calculated as previously described: The Cornell Voltage is calculated as the sum of the S wave in lead V3 + the R wave in lead aVL [15]. The Sokolow-Lyon LVH criteria is calculated as the sum of the S wave in lead V1 and the maximum of the R wave in either lead V5 or V6 [16].

**VCG Loop Morphology and Singular Value Decomposition:**

The singular value decomposition (SVD) is used to find the best fit planes for the QRS and T loops separately. Let $X' = X - c_x$, $Y' = Y - c_y$, and $Z' = Z - c_z$ where the centroid $\mathbf{c} = (c_x, c_y, c_z)$ is the mean QRS or T vector. Then let the matrix $\mathbf{M}'$ have the centroid-subtracted leads as columns. The SVD of $\mathbf{M}'$ is:

$$\mathbf{M}' = \mathbf{U}\mathbf{S}\mathbf{V}^T \tag{22}$$

After SVD, unit vectors that span the best fit plane for the VCG loop are given by the columns of $\mathbf{V}$.

The 3 singular values along the diagonal of matrix $\mathbf{S}$, when squared, give the proportion of mean-squared error or variance ($\sigma^2$) in the direction of the 3 corresponding basis vectors in matrix $\mathbf{V}$:

$$S = \begin{bmatrix} S_1 & 0 & 0 \\ 0 & S_2 & 0 \\ 0 & 0 & S_3 \end{bmatrix} \qquad \sigma^2 = \begin{bmatrix} S_1^2 & 0 & 0 \\ 0 & S_2^2 & 0 \\ 0 & 0 & S_3^2 \end{bmatrix} \tag{23}$$

The columns are arranged such that $S_1 > S_2 > S_3$. Assuming that the VCG loop is approximately planar, the third column of matrix $\mathbf{V}$ is normal to the best fit plane; let this vector be denoted by $\mathbf{n}$. Note that if the the VCG loop is perfectly coplanar, then $S_3 = 0$. The major and minor axes of the VCG loop are then given by the first and second columns of $\mathbf{V}$, respectively.

The degree of VCG loop coplanarity is assessed in 2 ways. First, the mean standard error (MSE) in the direction normal to the best fit plane ($S_3^2$) is obtained from squaring the 3rd singular value as noted above. If $S_3^2 = 0$ then all points in the loop are coplanar. Coplanarity is also assessed by calculating RMSE for points in the VCG loop compared to the points projected onto the best fit plane:

For a given point in the QRS or T loops $\mathbf{s} = [s_x, s_y, s_z]$ let $\mathbf{s}' = \mathbf{s} - \mathbf{c}$. Then the projection $s'_{\text{proj}}$ relative to the centroid $\mathbf{c}$ onto the best-fit plane is found by subtracting out the component of the vector in the direction of the normal vector $\mathbf{n}$ defined above:

$$\mathbf{s'}_{\text{proj}} = \mathbf{s}' - (\mathbf{s}' \cdot \mathbf{n})\mathbf{n} \tag{24}$$

The contribution to the RMSE is then the square root of the mean distance between corresponding points $\mathbf{s}$ and $\mathbf{s}_{\text{proj}}$. Using this metric, if all points are coplanar, RMSE = 0.

**VCG Loop Dihedral Angle:**

VCG loop dihedral angle is defined as the 3-dimensional angle between the unit normal vectors that define the QRS loop best fit plane ($N_{\text{QRS}}$) and the T loop best fit plane ($N_{\text{T}}$). By convention, the dihedral angle is an acute angle:

$$\text{Dihedral Angle} = \arccos(|N_{\text{QRS}} \cdot N_{\text{T}}|) \tag{25}$$

**VCG Loop Roundness:**

Assuming that the MSE in the direction normal to the best fit plane should be relatively small compared to the MSE in the plane itself, the "roundness" ($R$) of the VCG loop is defined as the ratio of the largest to second largest singular values:

$$R = S_1/S_2 \tag{26}$$

If $R = 1$ the VCG loop is a perfect circle, and as the value of $R$ increases above 1 the loop is more oval or oblong.

**VCG Loop Perimeter:**

QRS and T loop perimeter are calculated as the length of the QRS or T loop projected into the best fit plane with the set of points defined as $M_{\text{proj}}$, and is analogous to the QRS and T loop length which is the length of the loop without projection (set of points defined as $M$). The perimeter of $M_{\text{proj}}$ is calculated using Matlab polyshapes.

**VCG Loop Area:**

QRS and T loop area are calculated as the area of the QRS or T loop projected into the best fit plane with the set of points defined as $M_{\text{proj}}$. The area of $M_{\text{proj}}$ is calculated using Matlab polyshapes.

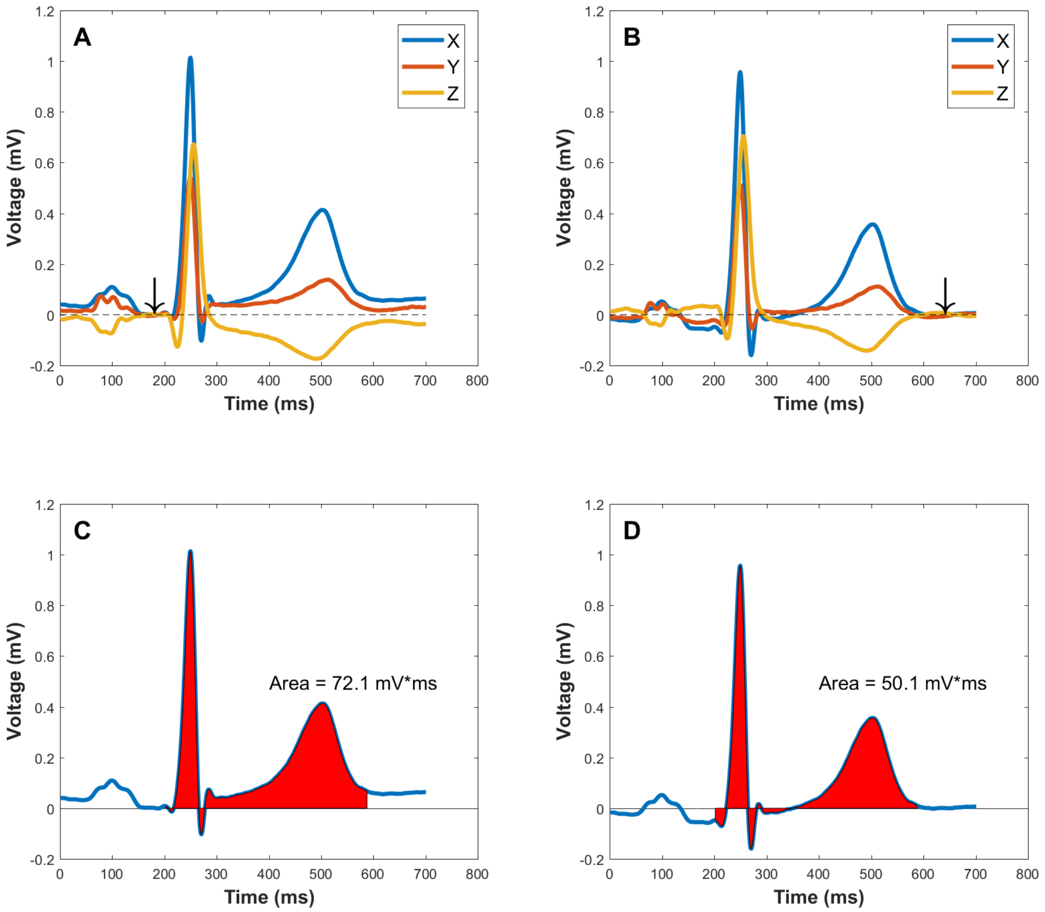


Supplemental Figure 1: Significant differences in QRST area calculations due to different definitions of the vectorcardiographic origin point. **A**: Median VCG beats (X, Y, Z) without any baseline processing where the onset of the QRS complex (arrow) is defined as zero voltage. **B**: After baseline correction using the method described in the text, the TP segment, a physiologically isoelectric interval, is now set as the zero voltage (arrow). **C** and **D**: Physiological baseline correction results in a 30% decrease in area under the X median beat. Reproduced with permission from [17].

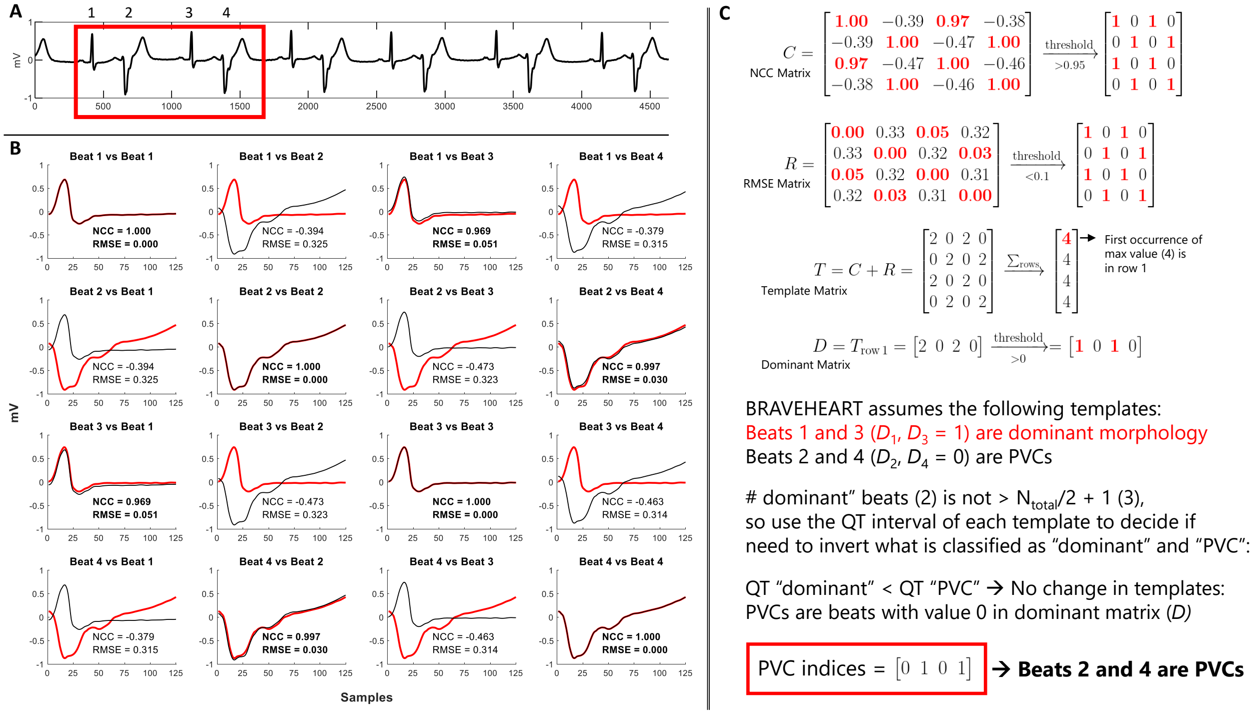


Supplemental Figure 2: Example of how the BRAVEHEART PVC detection algorithm works on an ECG with bigeminy and equal numbers of sinus beats and PVCs. **A:** A subset of 4 beats (2 sinus and 2 PVCs) are analyzed for illustrative purposes, but the algorithm would have the same result if all 12 beats were analyzed. **B:** After trimming, each beat is aligned with the remaining beats, and normalized cross correlation (NCC) and root mean squared error (RMSE) are calculated. **C:** Values of NCC and RMSE are placed in matrices $C$ and $R$, respectively, and then thresholds are applied as described in the text. The template matrix $T$ is formed by adding $C + R$ and summing the rows. The dominant matrix $D$ is formed by taking the first row of $T$ that has the maximum value of $T$, and then performing additional thresholding. BRAVEHEART starts by assuming that values of 1 in $D$ are the dominant morphology and values of 0 in $D$ are PVCs. In this case, the number of beats with dominant morphology is not $> n/2 + 1$, so the algorithm has to look at the QRS and QT interval of each morphology to decide if the "dominant" morphology is sinus or a PVC. In this case the QRS duration of the "dominant" morphology is <90% the QRS duration of the "PVC" morphology, so the template associated with the "dominant" morphology (narrow beats) is the true dominant morphology. The indices of PVCs are found by taking values of 0 in matrix $D$ (beats 2 and 4).
**Abbreviations**: PVC - premature ventricular contraction

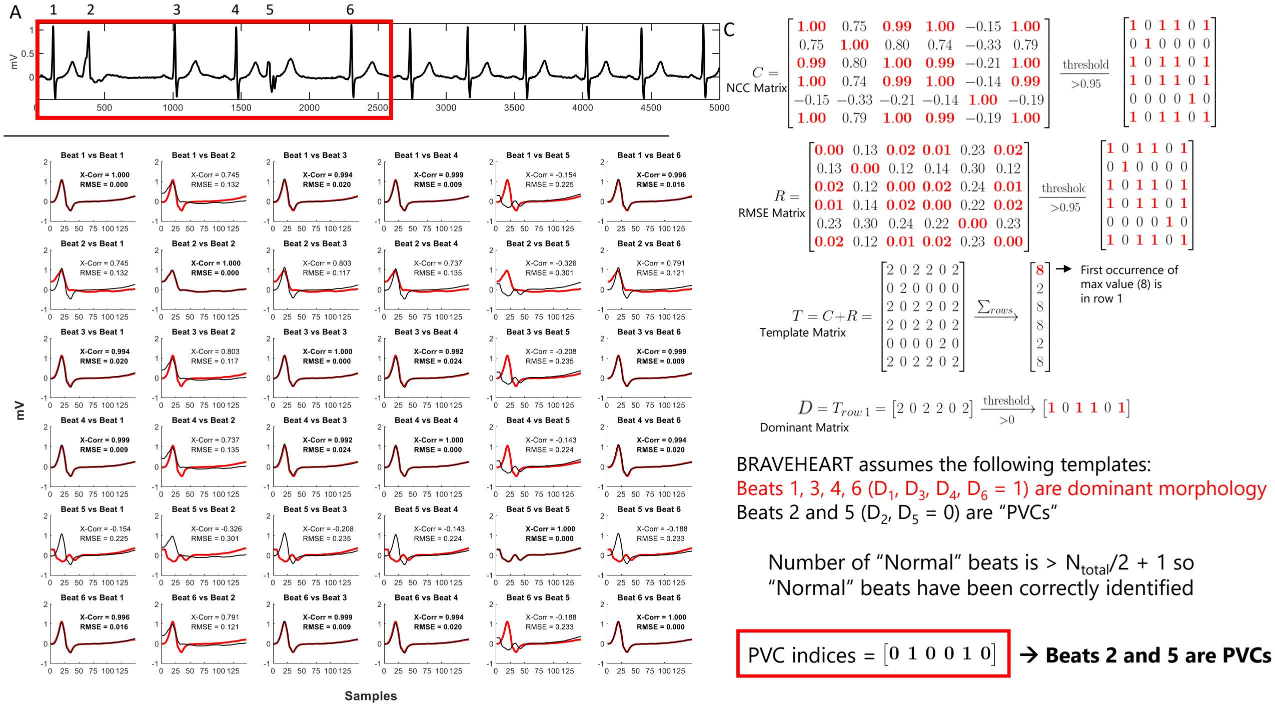


Supplemental Figure 3: Example of how the BRAVEHEART PVC detection algorithm works on an ECG with multiple PVC morphologies. **A:** A subset of 6 beats (4 sinus and 2 PVCs of different morphology) are analyzed for illustrative purposes, but the algorithm would have the same result if all 12 beats were analyzed. **B:** After trimming, each beat is aligned with the remaining beats, and normalized cross correlation (NCC) and root mean squared error (RMSE) are calculated. **C:** Values of NCC and RMSE are placed in matrices $C$ and $R$, respectively, and then thresholds are applied as described in the text. The template matrix $T$ is formed by adding $C + R$ and summing the rows. The dominant matrix $D$ is formed by taking the first row of $T$ that has the maximum value of $T$, and then performing additional thresholding. BRAVEHEART starts by assuming that values of 1 in $D$ are the dominant morphology and values of 0 in $D$ are PVCs. In this case, since the number of beats with dominant morphology is $> n/2 + 1$, the algorithm is complete and the indices of PVCs are found by taking values of 0 in matrix $D$ (beats 2 and 5).

**Abbreviations**: PVC - premature ventricular contraction

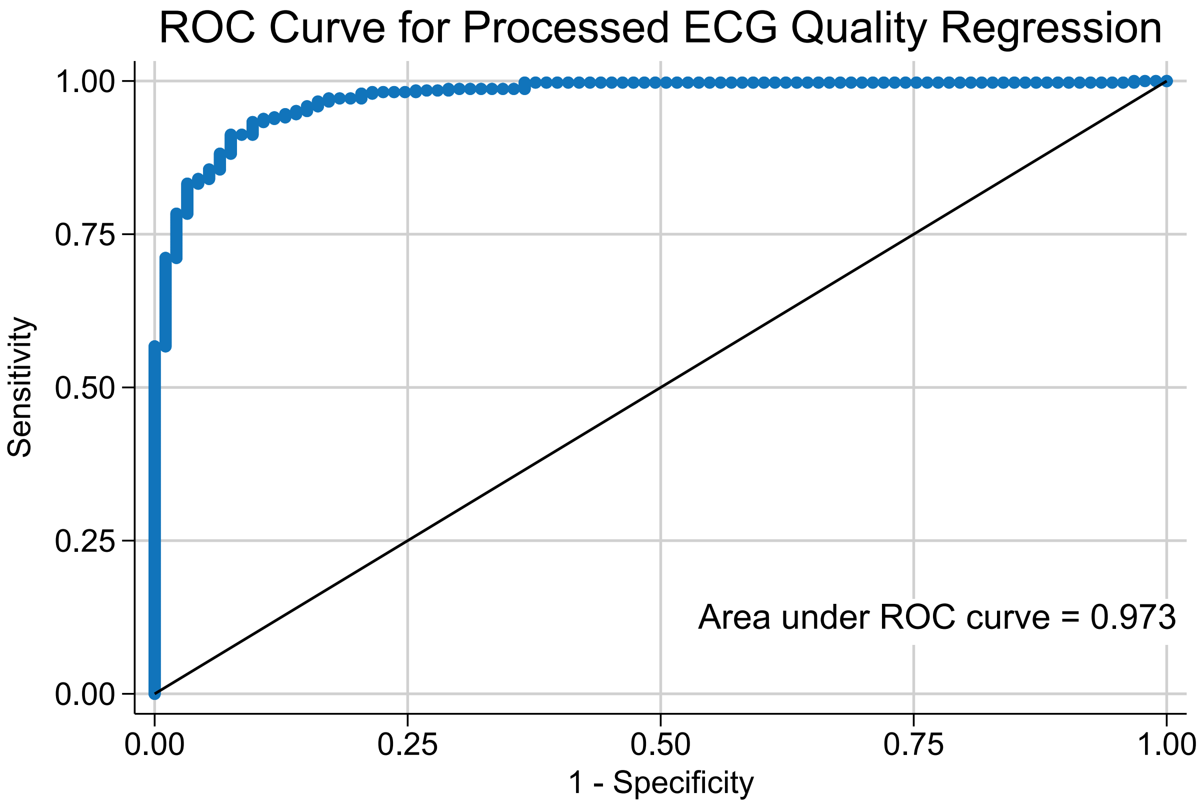


Supplemental Figure 4: Receiver Operating Curve for quality regression. The area under the receiver operating curve of 0.973 indicates that the final model has excellent predictive ability.

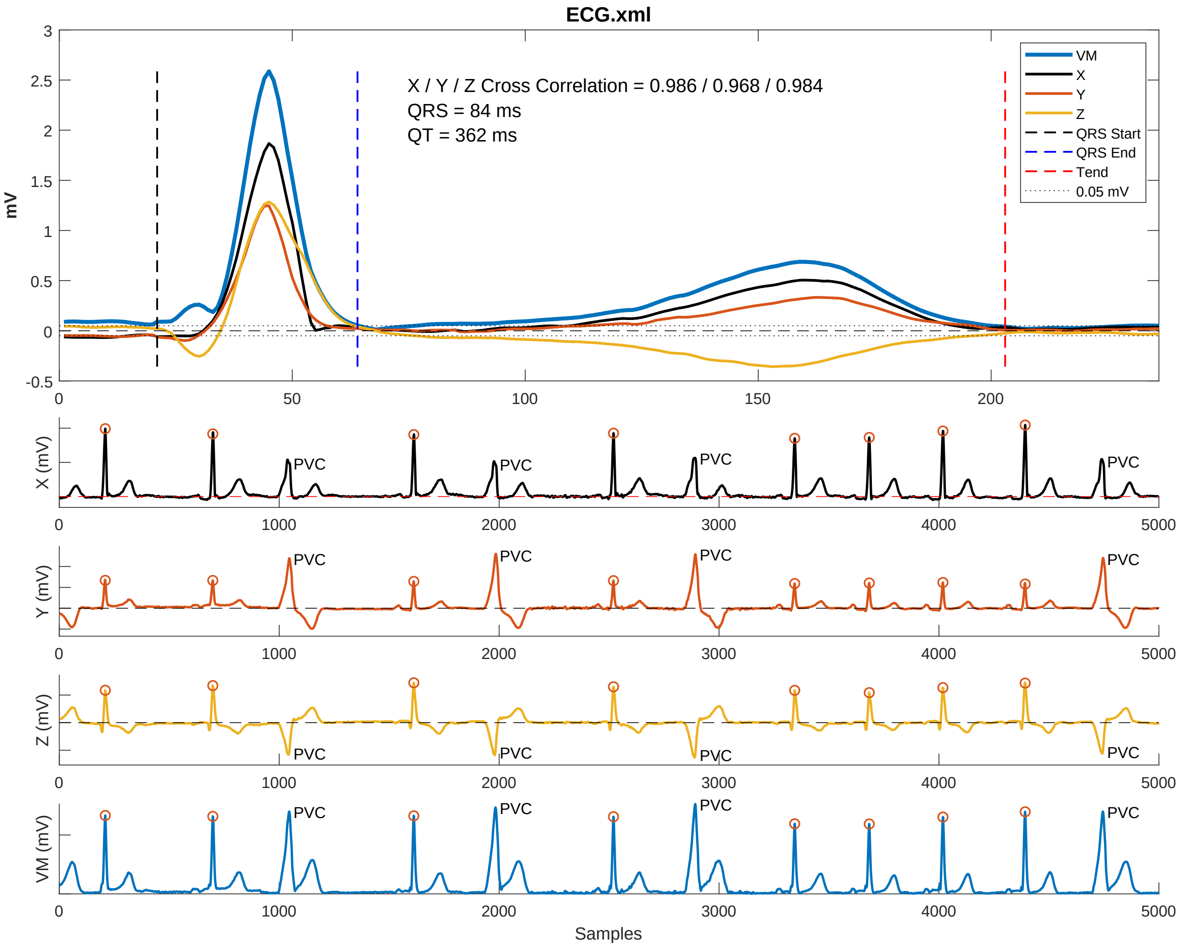


Supplemental Figure 5: Example of summary figure output. The X, Y, Z, and VM median beats and their annotations are show in the top panel, and the full VCG is shown in the bottom half of the figure. Beats included in the median beat have an orange circle at their R peak. In this example, beats without an orange circle are labeled as "PVC" to indicate they were detected as PVCs and removed from the median beat analysis. Cross correlation represents the average normalized cross correlation between all pairs of beats that make up the median beat and represents the quality of median beat construction with values very close to 1 indicating excellent beat alignment.

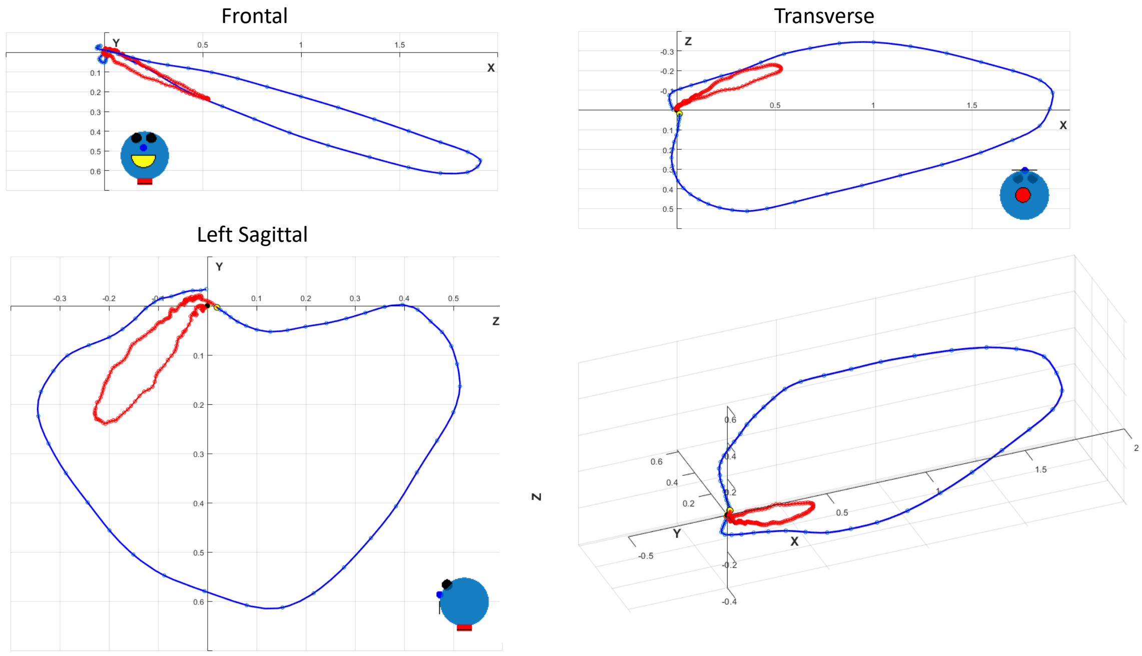


Supplemental Figure 6: Example of VCG generated from an ECG. The QRS loop is blue and the T wave loop is red.

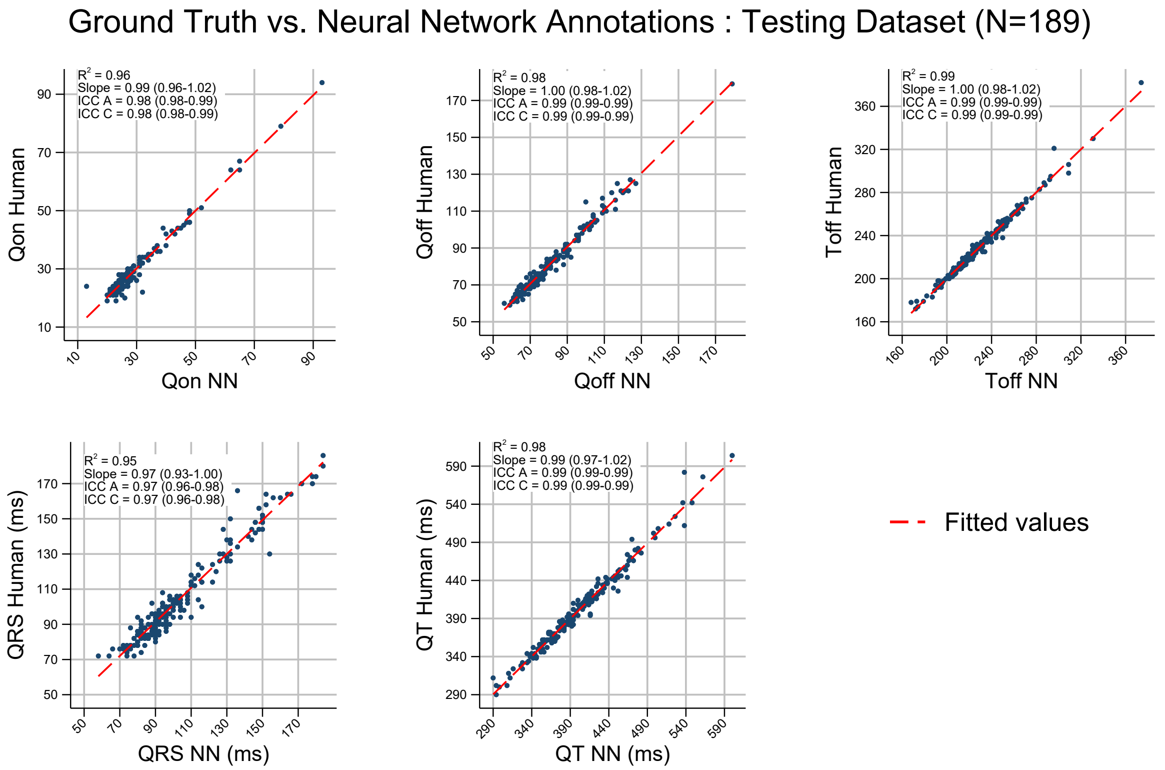


Supplemental Figure 7: Correlations between NN predictions and ground truth fiducial points for the initial 189 ECG testing dataset. Note that relationship between NN predicted and ground truth values of all fidicual points are very liner and ICC values are all very close to 1.

**Abbreviations**: ICC A - absolute agreement intraclass correlation coefficient, ICC C - consistency of agreement intraclass correlation coefficient.

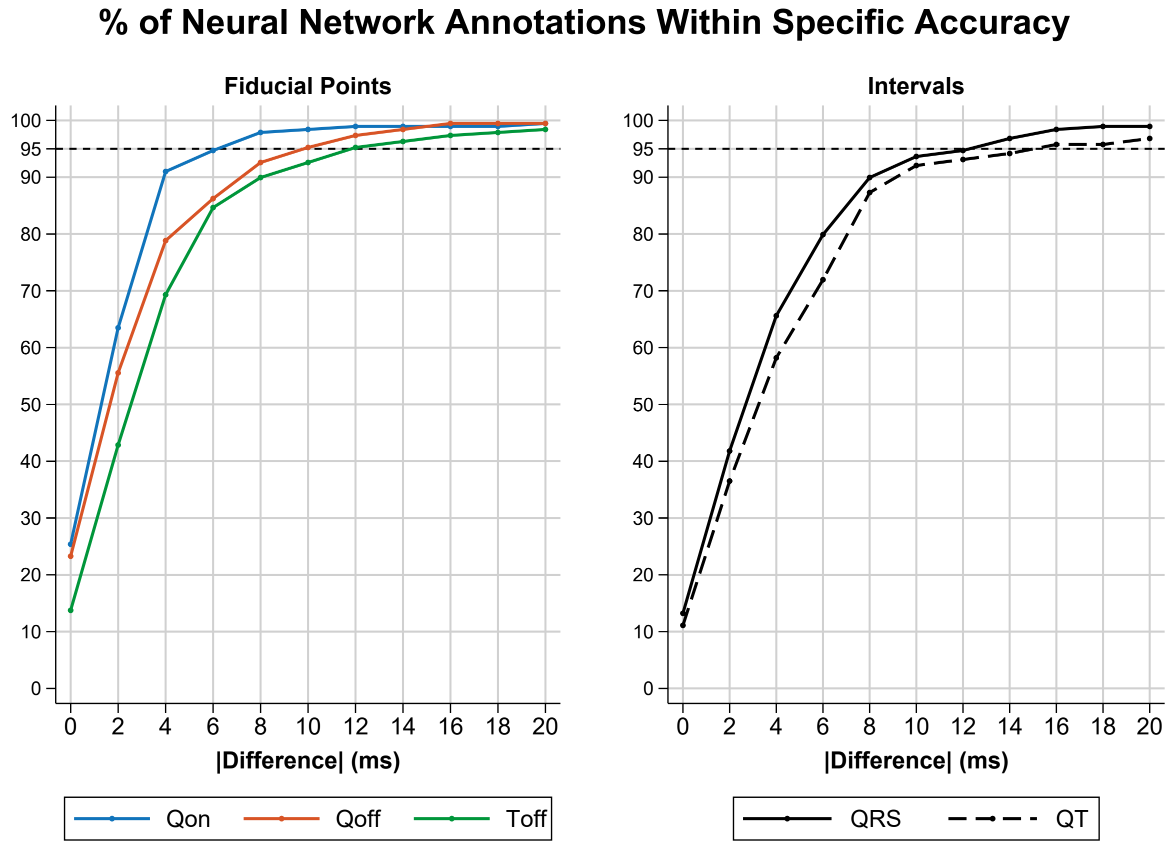


Supplemental Figure 8: Percentage of NN detected fiducial points/intervals that were within various errors (in ms) compared to ground truth for the initial 189 ECG testing dataset.

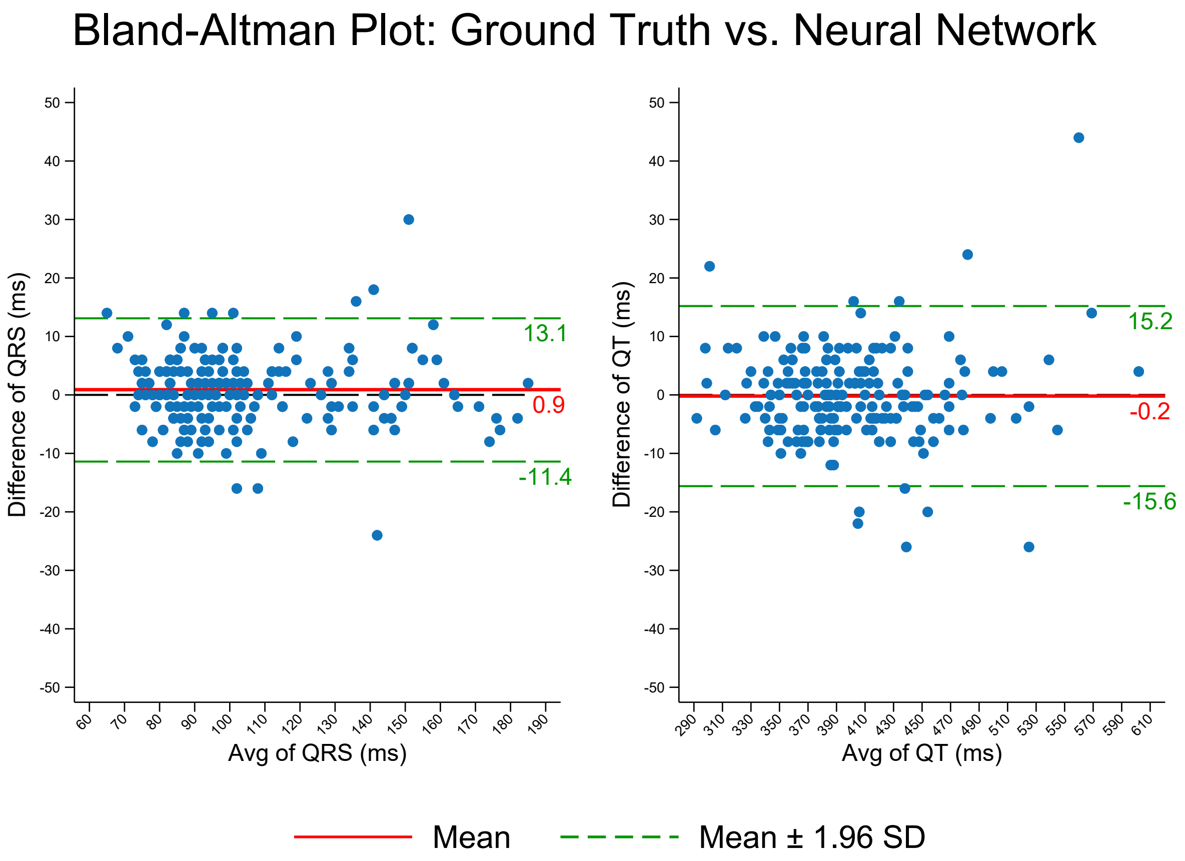


Supplemental Figure 9: Bland-Altman plot for QRS duration and QT interval for the initial 189 ECG testing dataset.

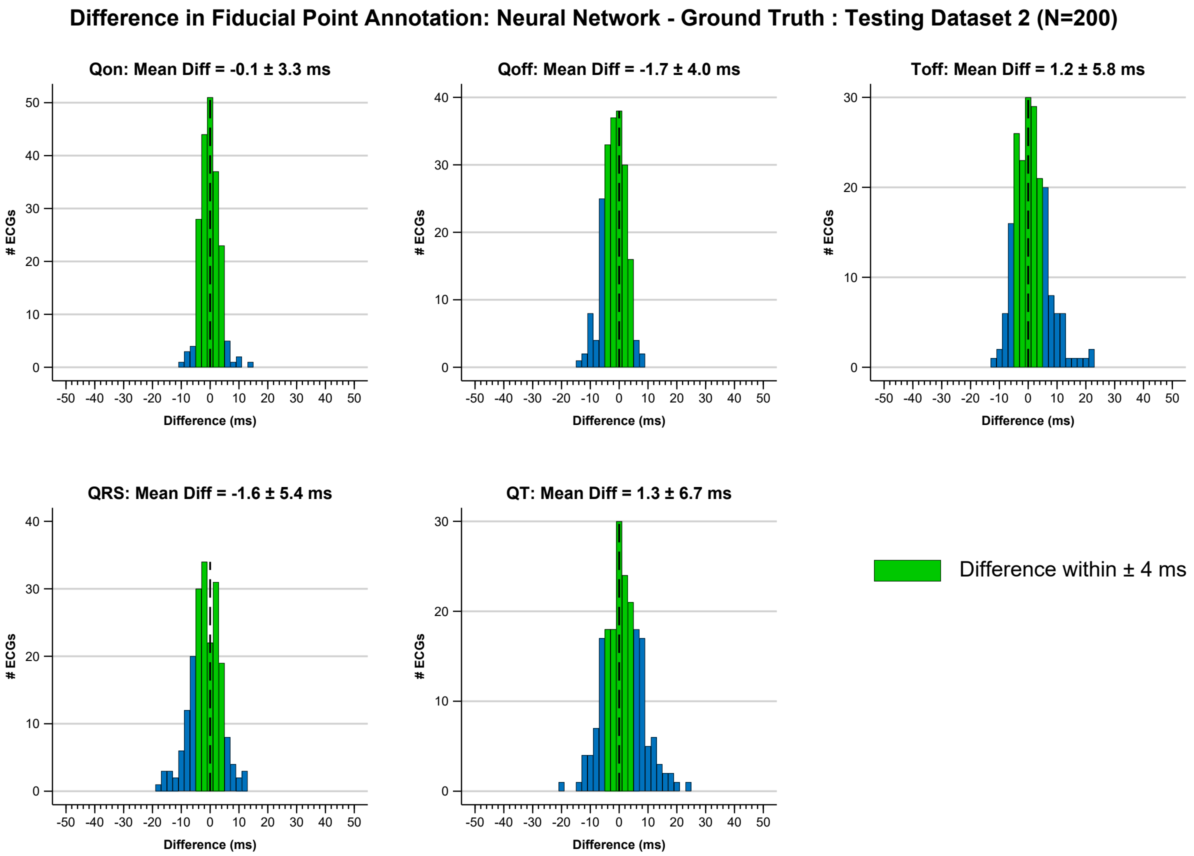


Supplemental Figure 10: Mean and standard deviation and histograms for results of NN testing for the second 200 ECG testing dataset. Bins where the difference between NN predictions and ground truth fiducial points were $\leq \pm$ 4 ms are shown in green.

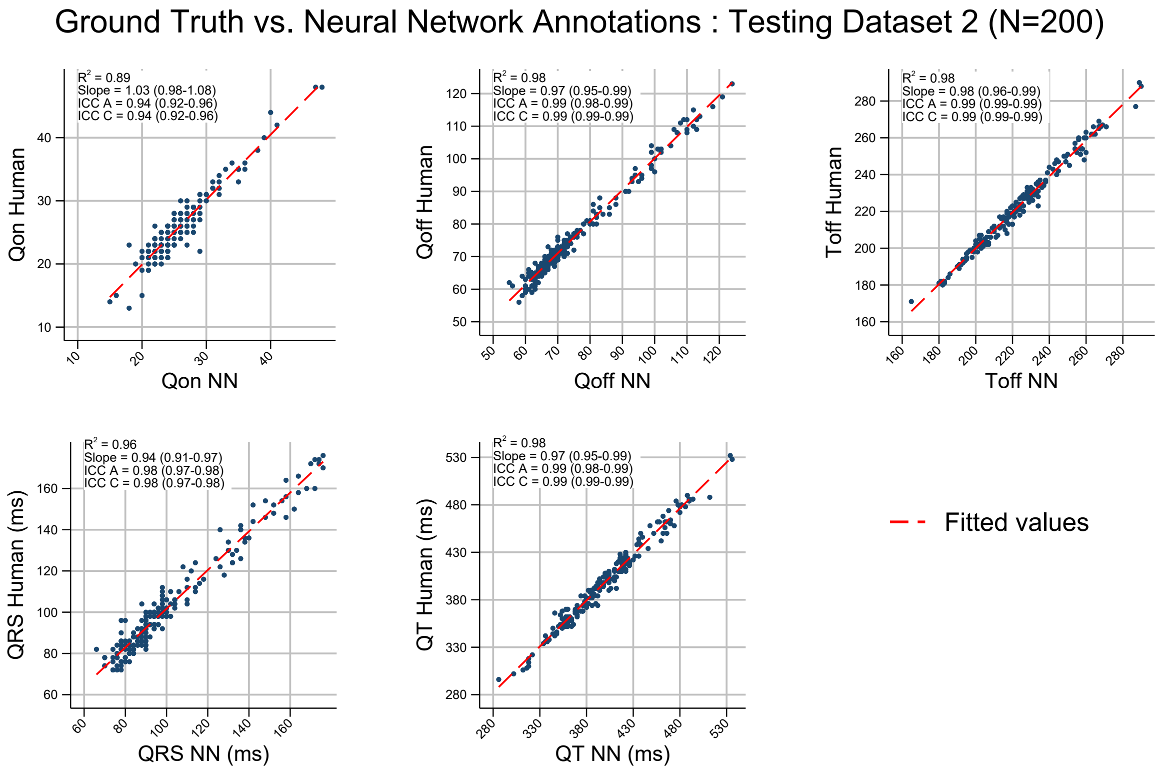


Supplemental Figure 11: Correlations between NN predictions and ground truth fiducial points for the second 200 ECG testing dataset. Note that relationship between NN predicted and ground truth values of all fidicual points are very liner and ICC values are all very close to 1.

**Abbreviations**: ICC A - absolute agreement intraclass correlation coefficient, ICC C - consistency of agreement intraclass correlation coefficient.

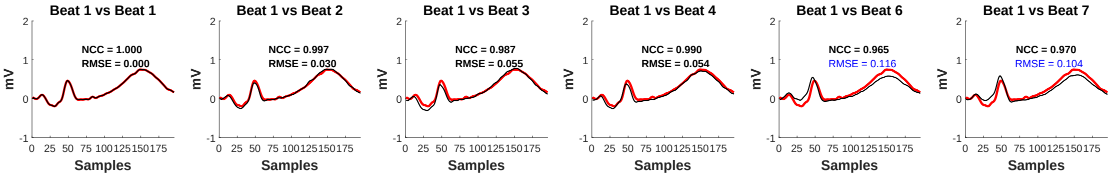


Supplemental Figure 12: Example where use of both NCC and RMSE helps prevent over-calling beats with slightly differnet morphology due to artifact/respiration as PVCs. In this case the RMSE and NCC thresholds were set to the nominal values of 0.1 and 95%, respectively. Beats 6 and 7 have RMSE just above the threshold, but have good NCC and should not be detected as PVCs.

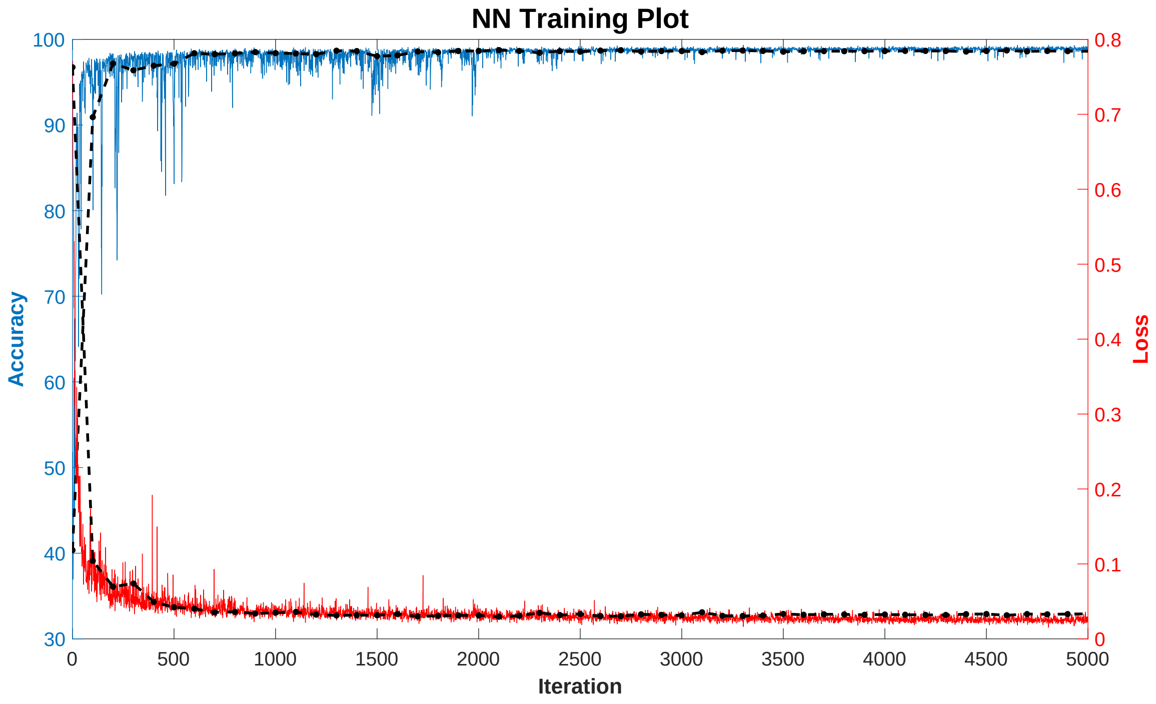


Supplemental Figure 13: Neural network training curves for the neural network included with BRAVEHEART for median bean annotation. The blue line represents training accuracy and the red line represents training loss. The dashed lines respresents validation training/loss.

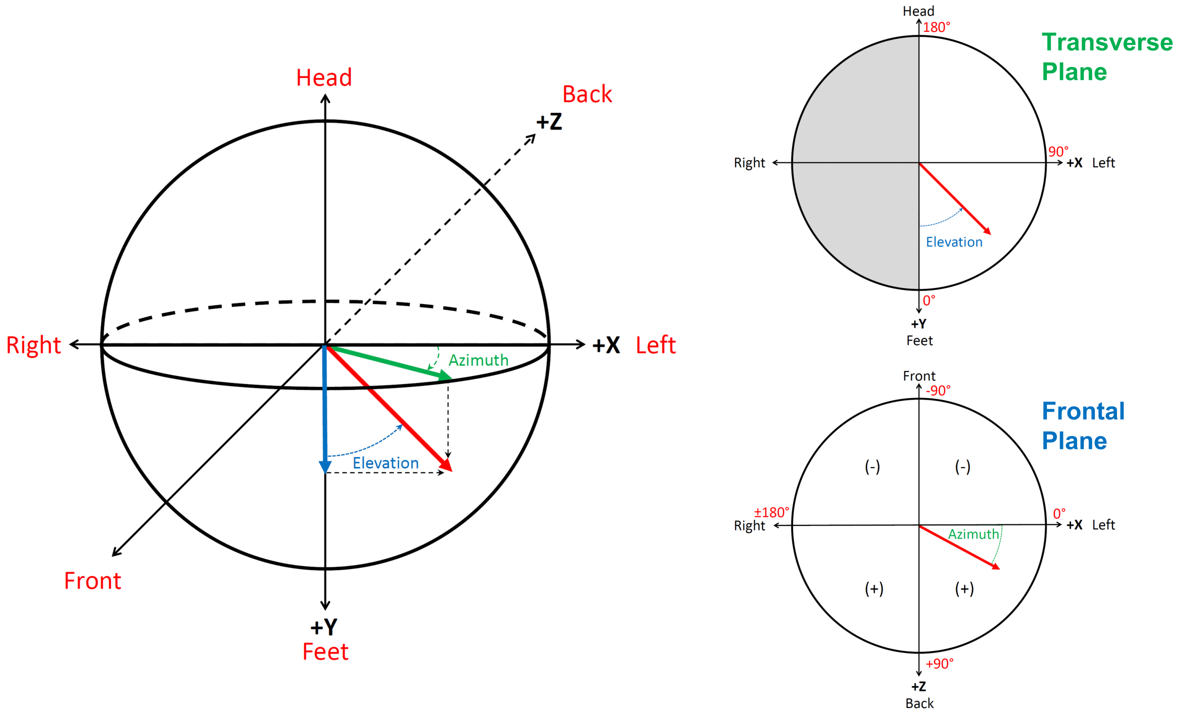


Supplemental Figure 14: Definitions of azimuth and elevation. A vector is shown in red. Elevation is the angle in the XY plane with range 0 to 180° with 0° defined as pointing towards the feet (+Y), and 180° pointing towards the head (-Y). Azimuth is the angle in the XZ plane with range 0 to ±180°. Azimuth 0° points towards the left (+X), while ±180° points towards the left (-X), with positive angles oriented posteriorly and negative angles oriented anteriorly.

Supplemental Table 1: Output Variables 1 of 3

| Variable | Description | Units |
|---|---|---|
| **Standard Intervals** | | |
| qrs_int | QRS duration | ms |
| qt_int | QT interval | ms |
| **QRS Area Vector** | | |
| XQ_area | Area under median X QRS complex | mV·ms |
| YQ_area | Area under median Y QRS complex | mV·ms |
| ZQ_area | Area under median Z QRS complex | mV·ms |
| q_area_mag | Magnitude of QRS area vector ([XQ_area, YQ_area, ZQ_area]) | mV·ms |
| q_area_az | Azimuth of QRS area vector | deg |
| q_area_el | Elevation of QRS area vector | deg |
| **T Area Vector** | | |
| XT_area | Area under median X T wave | mV·ms |
| YT_area | Area under median Y T wave | mV·ms |
| ZT_area | Area under median Z T wave | mV·ms |
| t_area_mag | Magnitude of T-wave area vector [XT_area, YT_area, ZT_area] | mV·ms |
| t_area_az | Azimuth of T-wave area vector | deg |
| t_area_el | Elevation of T-wave area vector | deg |
| **SVG** | | |
| svg_x | X component of SVG = XQ_area + XT_area | mV·ms |
| svg_y | Y component of SVG = YQ_area + YT_area | mV·ms |
| svg_z | Z component of SVG = ZQ_area + ZT_area | mV·ms |
| svg_area_mag | Magnitude of the SVG vector [svg_x, svg_z, svg_z] | mV·ms |
| svg_area_az | Azimuth of the SVG vector | deg |
| svg_area_el | Elevation of the SVG vector | deg |
| **SAI QRST** | | |
| sai_x | Area under the absolute value of the median X QRST complex | mV·ms |
| sai_y | Area under the absolute value of the median Y QRST complex | mV·ms |
| sai_z | Area under the absolute value of the median Z QRST complex | mV·ms |
| sai_qrst | SAI QRST = sai_x + sai_y + sai_z | mV·ms |
| sai_vm | Area under the absolute value of the median VM QRST complex | mV·ms |
| **QRS Peak Vector** | | |
| XQ_peak | Value of median X QRS complex at time of maximum distance from origin | mV |
| YQ_peak | Value of median Y QRS complex at time of maximum distance from origin | mV |
| ZQ_peak | Value of median Z QRS complex at time of maximum distance from origin | mV |
| q_peak_mag | Magnitude of peak QRS vector | mV |
| q_peak_az | Azimuth of peak QRS vector | mV |
| q_peak_el | Elevation of peak QRS vector | mV |
| **T Peak Vector** | | |
| XT_peak | Value of median X T wave at time of maximum distance from origin | mV |
| YT_peak | Value of median Y T wave at time of maximum distance from origin | mV |
| ZT_peak | Value of median Z T wave at time of maximum distance from origin | mV |
| t_peak_mag | Magnitude of peak T wave vector | mV |
| t_peak_az | Azimuth of peak T wave vector | mV |
| t_peak_el | Elevation of peak T wave vector | mV |
| **QRST Angles** | | |
| qrst_angle_area | Area QRST angle: 3D angle between area QRS and area T wave vectors | deg |
| qrst_angle_peak | Peak QRST angle:3D angle between peak QRS and peak T wave vectors | deg |
| qrst_angle_peak_frontal | Projection of area QRST angle into frontal plane | deg |
| qrst_angle_area_frontal | Projection of peak QRST angle into frontal plane | deg |
| TCRT | Total Cosine R to T (TCRT) | mV |
| TCRT_angle | TCRT Angle = acos(TCRT) | deg |

Supplemental Table 2: Output Variables 2 of 3

| Measurement | Description | Units |
|---|---|---|
| **LVH** | | |
| cornell_lvh_mv | Cornell LVH voltage | mV |
| sokolow_lvh_mv | Sokolow-Lyon LVH voltage | mV |
| **VCG Loop Length** | | |
| vcg_length_qrs | Length of QRS VCG loop | mV |
| vcg_length_t | Length of T wave VCG loop | mV |
| vcg_length_qrst | Length of QRST VCG loop = vcg_length_qrs + vcg_length_t | mV |
| **VCG Loop Speed** | | |
| speed_max | Maximum speed across the entire VCG loop | mV/ms |
| speed_min | Minimum speed across the entire VCG loop | mV/ms |
| speed_med | Median speed across the entire VCG loop | mV/ms |
| time_speed_max | Time after QRS onset of maximum VCG speed | ms |
| time_speed_min | Time after QRS onset of minimum VCG speed | ms |
| speed_qrs_max | Maximum speed across the QRS VCG loop | mV/ms |
| speed_qrs_min | Minimum speed across the QRS VCG loop | mV/ms |
| speed_qrs_med | Median speed across the QRS VCG loop | mV/ms |
| time_speed_qrs_max | Time after QRS onset of maximum QRS speed | ms |
| time_speed_qrs_min | Time after QRS onset of minimum QRS speed | ms |
| speed_t_max | Maximum speed across the T wave loop | mV/ms |
| speed_t_min | Minimum speed across the T wave loop | mV/ms |
| speed_t_med | Median speed across the T wave loop | mV/ms |
| time_speed_t_max | Time after QRS onset of maximum T-wave speed | ms |
| time_speed_t_min | Time after QRS onset of minimum T-wave speed | ms |
| **T Wave Morphology** | | |
| vm_tpeak_time | Time after QRS onset of peak of median VM Twave | ms |
| vm_tpeak_tend_abs_diff | Time difference between T wave peak and T wave end in median VM lead | ms |
| vm_tpeak_tend_ratio | Ratio between time of T wave peak and time of T wave end in median VM lead | – |
| **Lead Morphology** | [lead] is any of the 16 leads (I, II, III, avL, avR, avF, V1-V6, X, Y, Z, VM) | |
| [lead]_r_wave | Magnitude of R wave on median beat of [lead] | mV |
| [lead]_s_wave | Magnitude of S wave on median beat of [lead] | mV |
| [lead]_rs_wave | Magnitude of entire QRS complex = [lead]_r_wave + abs([lead]_s_wave | mV |
| [lead]_rs_ratio | Ratio of R wave to magnitude of entire QRS complex = [lead]_r_wave / [lead]_rs_wave | – |
| [lead]_sr_ratio | Ratio of S wave to magnitude of entire QRS complex = [lead]_s_wave / [lead]_rs_wave | – |
| [lead]_t_max | Maximum magnitude of T wave in [lead] | mV |
| [lead]_t_max_loc | Timing of T wave maximum (after QRS onset) in [lead] | ms |
| **VCG Loop Morphology** | | |
| qrsloop_residual | SVD variance from fitting QRS loop to a plane = qrs_S3$^2$ (0 = perfect fit) | – |
| qrsloop_rmse | RMSE for fit of QRS loop to best fit plane (0 = perfect fit) | mV |
| qrsloop_roundness | QRS loop roundness. 1 = perfect circle, larger values are increasingly elliptical | – |
| qrsloop_area | Area of QRS loop | mV |
| qrsloop_perimeter | Length of QRS loop projected into best fit plane | mV$^2$ |
| tloop_residual | SVD variance from fitting T loop to a plane = t_S3$^2$ (0 = perfect fit) | – |
| tloop_rmse | RMSE for fit of T loop to best fit plane (0 = perfect fit) | mV |
| tloop_roundness | T loop roundness. 1 = perfect circle, larger values are increasingly elliptical | – |
| tloop_area | Area of T loop | mV |
| tloop_perimeter | Length of T loop projected into best fit plane | mV$^2$ |
| qrs_loop_normal | Unit vector normal to best fit QRS loop plane | – |
| t_loop_normal | Unit vector normal to best fit T loop plane | – |
| qrst_dihedral_ang | Dihedral angle between best fit QRS loop and T loop planes | deg |

Supplemental Table 3: Output Variables 3 of 3

| Measurement | Description | Units |
|---|---|---|
| qrs_S1 | 1st singular value of QRS loop | – |
| qrs_S2 | 2nd singular value of QRS loop | – |
| qrs_S3 | 3rd singular value of QRS loop | – |
| t_S1 | 1st singular value of T loop | – |
| t_S2 | 2nd singular value of T loop | – |
| t_S3 | 3rd singular value of T loop | – |
| qrs_var_s1_total | % of total variance made up by 1st QRS singular value | % |
| qrs_var_s2_total | % of total variance made up by 2nd QRS singular value | % |
| qrs_var_s3_total | % of total variance made up by 3rd QRS singular value | % |
| t_var_s1_total | % of total variance made up by 1st T singular value | % |
| t_var_s2_total | % of total variance made up by 2nd T singular value | % |
| t_var_s3_total | % of total variance made up by 3rd T singular value | % |

Supplemental Table 4: Results of assessing PVC detection efficacy at different cutoffs for NCC and RMSE.

| NCC | RMSE | F1 | Acc | NPV | PPV | Spec | Sens | % Missed PVC |
|---|---|---|---|---|---|---|---|---|
| 80 | 0.05 | 0.959 (0.931-0.979) | 0.988 (0.980-0.994) | 0.989 (0.983-0.994) | 0.983 (0.958-0.998) | 0.997 (0.993-1.000) | 0.935 (0.901-0.966) | 6.5 (3.4-9.9) |
| 85 | 0.05 | 0.967 (0.940-0.986) | 0.991 (0.983-0.996) | 0.993 (0.988-0.997) | 0.974 (0.948-0.992) | 0.996 (0.991-0.999) | 0.961 (0.928-0.983) | 3.9 (1.7-7.2) |
| 90 | 0.05 | 0.962 (0.938-0.981) | 0.989 (0.981-0.994) | 0.995 (0.990-0.999) | 0.952 (0.925-0.977) | 0.992 (0.987-0.996) | 0.972 (0.944-0.995) | 2.8 (0.5-5.6) |
| 95 | 0.05 | 0.892 (0.855-0.923) | 0.965 (0.952-0.976) | 0.997 (0.991-1.000) | 0.818 (0.769-0.865) | 0.963 (0.951-0.973) | 0.980 (0.950-1.000) | 2.0 (0.0-5.0) |
| 99 | 0.05 | 0.735 (0.686-0.780) | 0.901 (0.879-0.920) | 0.990 (0.981-0.996) | 0.602 (0.545-0.654) | 0.893 (0.872-0.912) | 0.945 (0.902-0.978) | 5.5 (2.2-9.8) |
| Off | 0.05 | 0.758 (0.710-0.804) | 0.911 (0.892-0.930) | 0.992 (0.984-0.998) | 0.628 (0.573-0.687) | 0.904 (0.884-0.923) | 0.955 (0.916-0.988) | 4.5 (1.2-8.4) |
| 80 | 0.1 | 0.963 (0.938-0.982) | 0.989 (0.982-0.995) | 0.990 (0.985-0.995) | 0.984 (0.960-0.998) | 0.997 (0.993-1.000) | 0.943 (0.912-0.970) | 5.7 (3.0-8.8) |
| 85 | 0.1 | 0.969 (0.943-0.987) | 0.991 (0.983-0.996) | 0.993 (0.988-0.997) | 0.980 (0.954-0.996) | 0.997 (0.992-0.999) | 0.959 (0.931-0.982) | 4.1 (1.8-6.9) |
| 90 | 0.1 | 0.976 (0.949-0.993) | 0.993 (0.985-0.998) | 0.996 (0.990-0.999) | 0.978 (0.951-0.996) | 0.996 (0.992-0.999) | 0.974 (0.944-0.994) | 2.6 (0.6-5.6) |
| **95** | **0.1** | **0.977 (0.952-0.994)** | **0.993 (0.986-0.998)** | **0.997 (0.992-1.000)** | **0.971 (0.945-0.992)** | **0.995 (0.990-0.999)** | **0.982 (0.955-1.000)** | **1.8 (0.0-4.5)** |
| **99** | **0.1** | **0.975 (0.949-0.993)** | **0.993 (0.985-0.998)** | **0.998 (0.994-1.000)** | **0.962 (0.932-0.986)** | **0.993 (0.988-0.998)** | **0.988 (0.963-1.000)** | **1.2 (0.0-3.7)** |
| **Off** | **0.1** | **0.978 (0.951-0.994)** | **0.993 (0.985-0.998)** | **0.998 (0.993-1.000)** | **0.969 (0.942-0.990)** | **0.995 (0.990-0.998)** | **0.986 (0.959-1.000)** | **1.4 (0.0-4.1)** |
| 80 | 1.5 | 0.935 (0.903-0.962) | 0.982 (0.973-0.989) | 0.982 (0.974-0.988) | 0.983 (0.957-1.000) | 0.997 (0.993-1.000) | 0.892 (0.849-0.932) | 10.8 (6.8-15.1) |
| 85 | 1.5 | 0.951 (0.921-0.974) | 0.986 (0.978-0.993) | 0.987 (0.979-0.992) | 0.983 (0.956-1.000) | 0.997 (0.993-1.000) | 0.921 (0.881-0.954) | 7.9 (4.6-11.9) |
| 90 | 1.5 | 0.963 (0.934-0.984) | 0.989 (0.981-0.995) | 0.990 (0.984-0.995) | 0.984 (0.958-1.000) | 0.997 (0.993-1.000) | 0.943 (0.907-0.973) | 5.7 (2.7-9.3) |
| 95 | 1.5 | 0.964 (0.934-0.984) | 0.990 (0.981-0.995) | 0.991 (0.984-0.996) | 0.984 (0.958-1.000) | 0.997 (0.993-1.000) | 0.945 (0.906-0.973) | 5.5 (2.7-9.4) |
| 99 | 1.5 | 0.966 (0.940-0.986) | 0.990 (0.983-0.996) | 0.991 (0.985-0.997) | 0.984 (0.958-1.000) | 0.997 (0.993-1.000) | 0.949 (0.915-0.979) | 5.1 (2.1-8.5) |
| Off | 1.5 | 0.964 (0.935-0.985) | 0.990 (0.981-0.996) | 0.991 (0.984-0.996) | 0.984 (0.958-1.000) | 0.997 (0.993-1.000) | 0.945 (0.908-0.977) | 5.5 (2.3-9.2) |
| 80 | Off | 0.954 (0.924-0.975) | 0.987 (0.979-0.993) | 0.988 (0.981-0.993) | 0.983 (0.957-0.998) | 0.997 (0.993-1.000) | 0.927 (0.888-0.958) | 7.3 (4.2-11.2) |
| 85 | Off | 0.968 (0.939-0.986) | 0.991 (0.982-0.996) | 0.993 (0.987-0.997) | 0.980 (0.953-0.996) | 0.997 (0.992-0.999) | 0.957 (0.924-0.980) | 4.3 (2.0-7.6) |
| 90 | Off | 0.972 (0.947-0.988) | 0.992 (0.984-0.997) | 0.996 (0.991-0.999) | 0.967 (0.943-0.987) | 0.994 (0.990-0.998) | 0.976 (0.948-0.996) | 2.4 (0.4-5.2) |
| 95 | Off | 0.902 (0.868-0.930) | 0.969 (0.958-0.978) | 0.996 (0.990-0.999) | 0.839 (0.792-0.880) | 0.968 (0.958-0.977) | 0.974 (0.943-0.996) | 2.6 (0.4-5.7) |
| 99 | Off | 0.512 (0.465-0.558) | 0.754 (0.723-0.783) | 0.975 (0.962-0.985) | 0.360 (0.318-0.404) | 0.731 (0.697-0.763) | 0.888 (0.833-0.933) | 11.2 (6.7-16.7) |

| ECG Diagnosis | Training N (%) | Testing N (%) | P Value |
|---|---|---|---|
| Sinus rhythm | 766 (80.9) | 154 (81.5) | 0.85 |
| Atrial fibrillation/flutter | 85 (9.0) | 19 (10.1) | 0.64 |
| Atrial pacing | 27 (2.9) | 5 (2.6) | 0.88 |
| Ventricular pacing | 94 (9.9) | 15 (7.9) | 0.4 |
| Left bundle branch block | 67 (7.1) | 6 (3.2) | 0.046 |
| Right bundle branch block | 72 (7.6) | 18 (9.5) | 0.37 |
| Nonspecific IVCD | 54 (5.7) | 9 (4.8) | 0.61 |
| Hemiblock | 48 (5.1) | 11 (5.8) | 0.67 |
| Bifascicular block | 27 (2.9) | 4 (2.1) | 0.57 |
| Left ventricular hypertrophy | 124 (13.1) | 20 (10.6) | 0.34 |
| Myocardial infarction | 20 (2.1) | 4 (2.1) | 1.0 |

Supplemental Table 5: ECG features used for neural network training (N = 947) and testing (N = 189). Overall features were well matched with the exception of a borderline lower rate of left bundle branch block in the testing dataset (p=0.46).

| ECG Diagnosis | N (%) |
|---|---|
| Sinus rhythm | 174 (87.0) |
| Atrial fibrillation/flutter | 16 (8.0) |
| Atrial pacing | 1 (0.5) |
| Ventricular pacing | 12 (6.0) |
| Left bundle branch block | 7 (3.5) |
| Right bundle branch block | 15 (7.5) |
| Nonspecific IVCD | 7 (3.5) |
| Hemiblock | 8 (4.01) |
| Bifascicular block | 2 (1.0) |
| Left ventricular hypertrophy | 20 (10.0) |
| Myocardial infarction | 1 (0.5) |

Supplemental Table 6: Diagnoses for the second 200 ECG neural network testing dataset.

[12] J. L. Willems, P. Arnaud, J. H. van Bemmel, P. J. Bourdillon, C. Brohet, S. Dalla Volta, J. D. Andersen, R. Degani, B. Denis, and M. Demeester. Assessment of the performance of

electrocardiographic computer programs with the use of a reference data base. *Circulation*, 71(3):523–534, Mar 1985.
